## Supplementary Information for "Risk mapping novel respiratory pathogens with large-scale dynamic contact networks"

#### Contents

**SI A: Sensitivity test of early epidemic to the number of seed actors** **2**

**SI B: Sensitivity of seed risk scores to time horizon length** **3**

**SI C: Sensitivity of transmission risk scores to time horizon length** **4**

**SI D: Effect of stochasticity for the impact of behavioural changes and targeted interventions** **26**

**SI E: Sensitivity of mobility restriction intervention to municipality population size** **27**

**SI F: Calculation of $\mathcal{R}_0$** **27**

### SI A: Sensitivity test of early epidemic to the number of seed actors

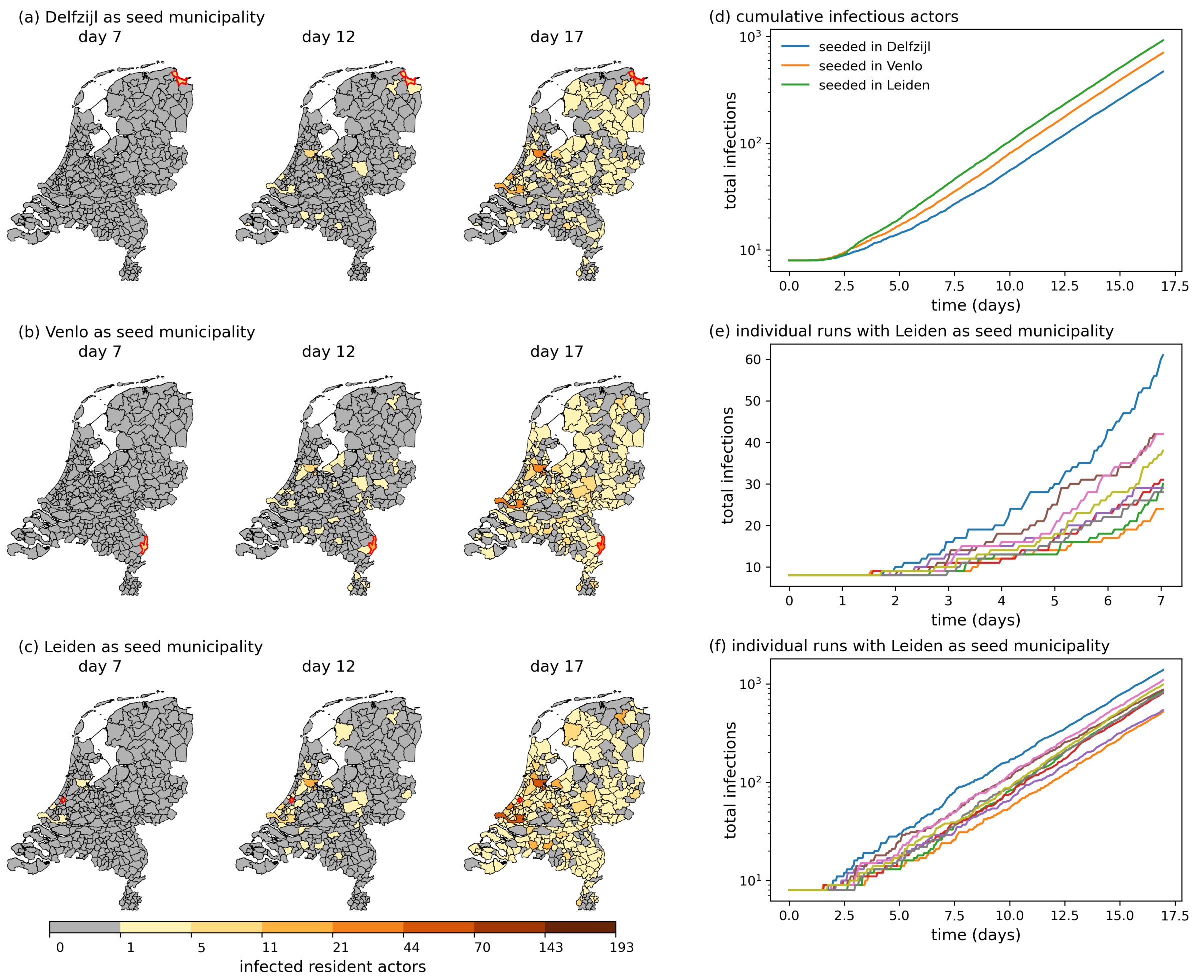


**Figure SI.1:** Reproduction of Fig. 2 of the main text with eight seed actors belonging to the working adults demographic group. The "infection intensity maps" of the Netherlands, i.e., cumulative number of infectious actors over 17 days, following pathogen introduction (seeding) amongst eight working adults of Delfzijl (a), Venlo (b), and Leiden (c) on day zero. For all three seed municipalities we also show the total cumulative infectious actors in panel (d) over the full 17 days. Compared to Fig. 2 of the main text, spatial spread occurs faster and the overall cumulative number of infectious actors is higher by a factor $\approx 8/5$, see SI Fig. SI.2.

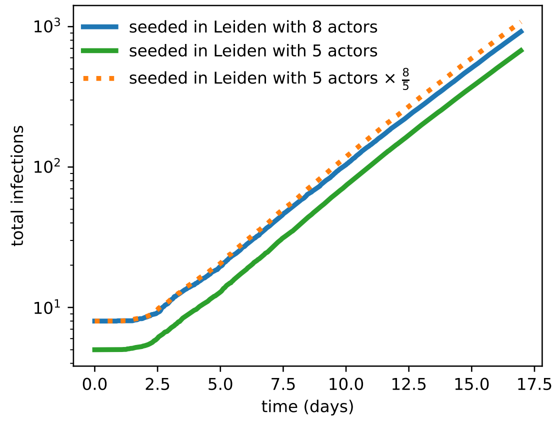


**Figure SI.2:** Comparison between the total number of infections in the Netherlands with five and eight actors, using Leiden as seed municipality for simulations with five seed actors. The total number of national infections is higher in the latter case by a factor $\approx 8/5$ at all times. This figure demonstrates robustness of our approach to scaling up the initial number of seed actors.

### SI B: Sensitivity of seed risk scores to time horizon length

We chose 17 day as the time horizon for calculating the seed and transmission risk scores in the main text. Here we test the sensitivity of the seed risk score by reproducing them for 14 (Fig. SI.3) and 21 days (Fig. SI.3). For visual comparison, we also reproduce Fig. 3 from the main text (Fig. SI.4).

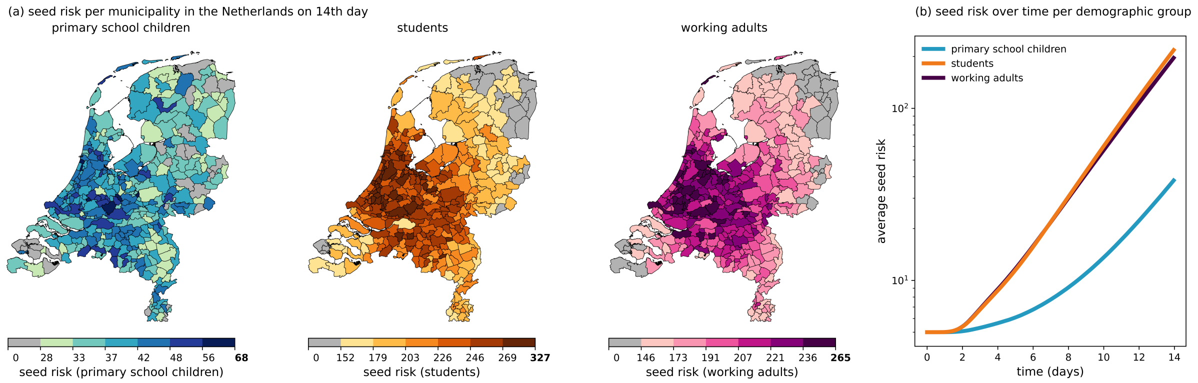


**Figure SI.3:** Seed risk maps (a) with 14 days as time horizon length. Seed risk averages over time per demographic group of seed actors shown in panel (b)

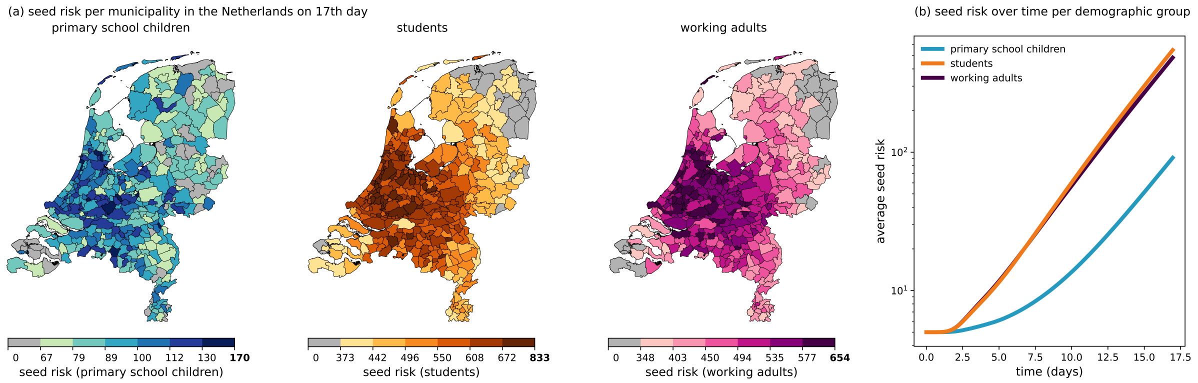


**Figure SI.4:** Seed risk maps (a) with 17 days as time horizon length. Seed risk averages over time per demographic group of seed actors shown in panel (b). This figure shows the same data as Fig. 3 in the main text.

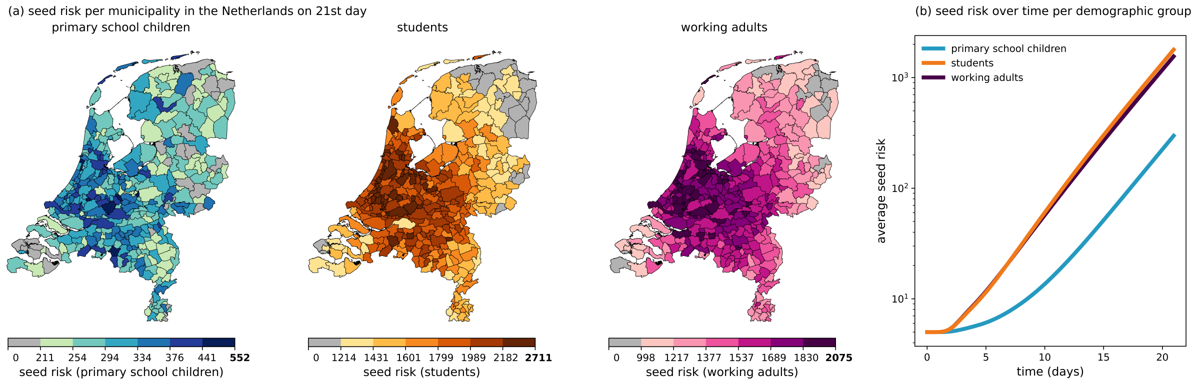


**Figure SI.5:** Seed risk maps (a) with 21 days as time horizon length. Seed risk averages over time per demographic group of seed actors shown in panel (b)

The seed risk scores for students at days 14, 17 and 21 can be found in the Supplementary Tabs. SI.1-SI.3 below.

Further, we also test the sensitivity of the seed risk score for the students demographic group to the time horizon by assigning all municipalities a rank based on their seed risk scores and plotting these ranks at 17 and 21 days days as function of the ranks at 14 days in Fig. SI.6.

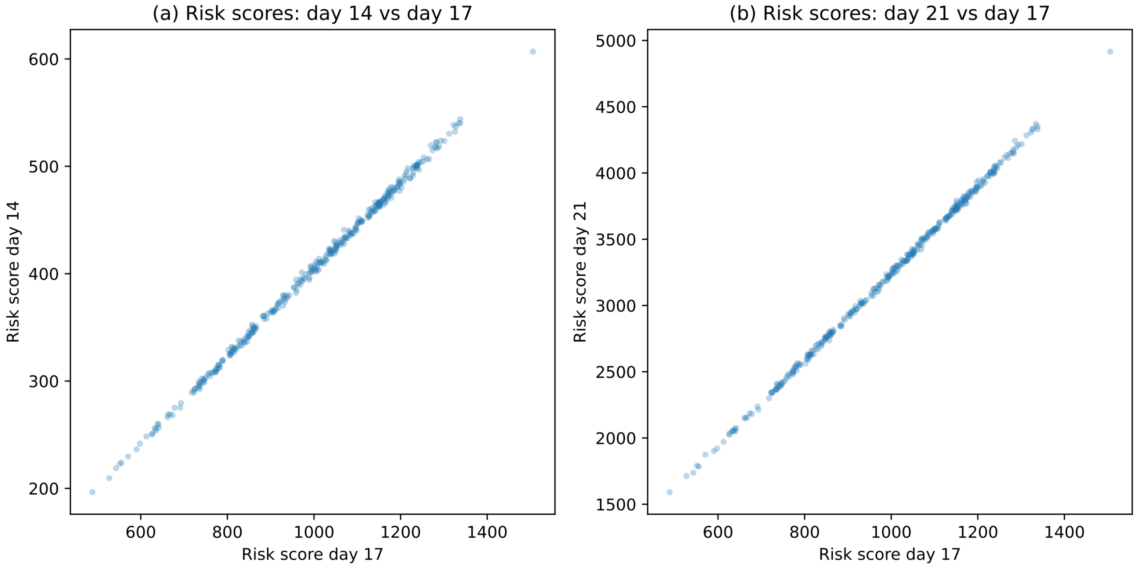


**Figure SI.6:** Scatter plots between the seed risk scores for the students demographic group between days 14, 17 and 21, for each municipality for seed risk scores at 14, 17, and 21 days as shown in the maps of Figs. SI.3, SI.4, SI.5, and Tabs. SI.1-SI.3. In panels (a-b) the day-pairs 14-17 days and 21-17 days are shown. These figures show that the risk scores are strongly correlated in their time horizons, indicated by the near-perfect visual linearity of the scatter plots.

.

### SI C: Sensitivity of transmission risk scores to time horizon length

**Supplementary Table SI.1:** The full list of population (data from 2019) and 14-days transmission risk scores and seed risk scores of the municipalities in the Netherlands after an initial introduction in students. See definition of transmission risk score in the main text.

| Municipality | Population size (×1000) | Population percentage | Percentage of transmissions | Transmission risk | (90% range) | Seed Risk | (90% range) |
|---|---|---|---|---|---|---|---|
| ’s-Gravenhage | 535.6 | 3.22% | 3.27% | 2.5 | 2.12-2.96 | 436.6 | 197-905 |
| ’s-Hertogenbosch | 153.5 | 0.92% | 0.95% | 2.5 | 1.99-3.27 | 426.7 | 168-751 |
| Aa en Hunze | 24.8 | 0.15% | 0.12% | 2.0 | 0.91-5.38 | 256.1 | 70-477 |
| Aalsmeer | 31.0 | 0.19% | 0.22% | 2.9 | 2.03-4.44 | 517.4 | 235-1,063 |
| Aalten | 26.5 | 0.16% | 0.12% | 1.9 | 0.87-3.30 | 260.1 | 65-612 |
| Achtkarspelen | 27.5 | 0.17% | 0.13% | 2.0 | 1.11-3.58 | 255.7 | 89-505 |
| Alblasserdam | 19.7 | 0.12% | 0.14% | 2.9 | 1.77-4.52 | 488.9 | 222-948 |
| Albrandswaard | 24.8 | 0.15% | 0.17% | 2.8 | 1.70-4.29 | 461.2 | 187-846 |
| Alkmaar | 108.2 | 0.65% | 0.63% | 2.4 | 1.81-2.99 | 431.9 | 169-825 |
| Almelo | 72.4 | 0.43% | 0.35% | 2.0 | 1.41-2.93 | 314.9 | 92-656 |
| Almere | 206.2 | 1.24% | 1.29% | 2.6 | 2.11-3.21 | 458.8 | 201-843 |
| Alphen aan den Rijn | 110.2 | 0.66% | 0.76% | 2.8 | 2.26-3.76 | 452.0 | 254-728 |
| Alphen-Chaam | 9.5 | 0.06% | 0.05% | 2.3 | 1.04-4.17 | 396.2 | 186-714 |
| Altena | 11.2 | 0.07% | 0.12% | 4.6 | 2.54-8.06 | 292.6 | 99-524 |
| Ameland | 3.2 | 0.02% | 0.01% | 1.6 | 0.31-3.99 | 291.5 | 70-640 |
| Amersfoort | 155.6 | 0.93% | 0.99% | 2.6 | 2.03-3.37 | 395.8 | 164-777 |
| Amstelveen | 90.1 | 0.54% | 0.57% | 2.6 | 1.95-3.22 | 447.5 | 190-786 |
| Amsterdam | 860.2 | 5.16% | 5.52% | 2.6 | 2.26-3.19 | 458.1 | 168-924 |
| Apeldoorn | 161.4 | 0.97% | 0.89% | 2.2 | 1.69-2.96 | 412.4 | 146-890 |
| Appingedam | 11.2 | 0.07% | 0.04% | 1.5 | 0.52-3.29 | 222.9 | 60-423 |
| Arnhem | 158.3 | 0.95% | 0.92% | 2.4 | 1.84-3.21 | 485.3 | 163-811 |
| Assen | 67.4 | 0.40% | 0.33% | 2.0 | 1.29-3.22 | 289.2 | 118-606 |
| Asten | 16.3 | 0.10% | 0.10% | 2.5 | 1.18-4.50 | 380.2 | 172-722 |
| Baarle-Nassau | 6.4 | 0.04% | 0.03% | 2.2 | 0.78-4.44 | 407.2 | 142-778 |
| Baarn | 24.2 | 0.15% | 0.17% | 2.9 | 1.89-4.61 | 463.0 | 161-788 |
| Barendrecht | 48.0 | 0.29% | 0.33% | 2.8 | 1.96-3.85 | 508.3 | 223-833 |
| Barneveld | 57.3 | 0.34% | 0.36% | 2.5 | 1.90-3.57 | 409.1 | 158-708 |
| Beek | 15.3 | 0.09% | 0.08% | 2.2 | 1.20-3.97 | 375.6 | 137-659 |
| Beekdaelen | 11.2 | 0.07% | 0.05% | 1.8 | 0.96-3.54 | 351.4 | 0-636 |
| Beemster | 9.2 | 0.06% | 0.06% | 2.6 | 1.26-4.45 | 488.3 | 225-905 |

Continued on next page

| Municipality | Population size (×1000) | Population percentage | Percentage of transmissions | Transmission risk | (90% range) | Seed Risk | (90% range) |
|---|---|---|---|---|---|---|---|
| Beesel | 13.0 | 0.08% | 0.06% | 2.0 | 0.91-4.05 | 336.6 | 113-649 |
| Berg en Dal | 34.3 | 0.21% | 0.23% | 2.8 | 1.60-3.96 | 421.4 | 155-847 |
| Bergeijk | 17.9 | 0.11% | 0.10% | 2.2 | 1.27-3.47 | 345.4 | 128-629 |
| Bergen (L.) | 12.6 | 0.08% | 0.07% | 2.4 | 1.17-3.89 | 325.9 | 113-682 |
| Bergen (NH.) | 29.4 | 0.18% | 0.16% | 2.2 | 1.31-3.25 | 370.1 | 133-657 |
| Bergen op Zoom | 66.1 | 0.40% | 0.37% | 2.3 | 1.45-4.54 | 307.8 | 83-590 |
| Berkelland | 43.4 | 0.26% | 0.23% | 2.2 | 1.43-3.17 | 349.3 | 115-754 |
| Bernheze | 30.3 | 0.18% | 0.21% | 2.8 | 1.71-4.18 | 433.3 | 213-686 |
| Best | 29.3 | 0.18% | 0.18% | 2.6 | 1.63-3.75 | 422.3 | 161-930 |
| Beuningen | 25.3 | 0.15% | 0.17% | 2.7 | 1.54-4.01 | 420.9 | 192-776 |
| Beverwijk | 40.6 | 0.24% | 0.26% | 2.6 | 1.90-3.79 | 463.3 | 188-993 |
| Bladel | 19.8 | 0.12% | 0.12% | 2.4 | 1.30-4.16 | 422.7 | 197-703 |
| Blaricum | 10.9 | 0.07% | 0.07% | 2.8 | 1.40-4.62 | 435.4 | 192-723 |
| Bloemendaal | 22.8 | 0.14% | 0.15% | 2.7 | 1.79-3.85 | 518.0 | 240-842 |
| Bodegraven-Reeuwijk | 33.9 | 0.20% | 0.25% | 3.0 | 2.00-3.83 | 500.6 | 238-882 |
| Boekel | 10.2 | 0.06% | 0.07% | 2.7 | 1.14-5.24 | 519.6 | 209-985 |
| Borger-Odoorn | 24.9 | 0.15% | 0.10% | 1.6 | 0.89-2.91 | 209.6 | 60-415 |
| Borne | 22.7 | 0.14% | 0.10% | 1.9 | 1.00-2.86 | 311.4 | 94-674 |
| Borsele | 22.2 | 0.13% | 0.11% | 2.0 | 1.10-3.15 | 348.9 | 85-727 |
| Boxmeer | 28.4 | 0.17% | 0.17% | 2.4 | 1.35-4.40 | 433.4 | 186-783 |
| Boxtel | 30.1 | 0.18% | 0.19% | 2.6 | 1.70-3.87 | 428.1 | 213-843 |
| Breda | 183.4 | 1.10% | 1.10% | 2.5 | 1.88-3.22 | 498.1 | 214-869 |
| Brielle | 16.9 | 0.10% | 0.09% | 2.3 | 1.29-4.09 | 440.3 | 192-887 |
| Bronckhorst | 35.7 | 0.21% | 0.18% | 2.1 | 1.32-3.13 | 349.4 | 152-668 |
| Brummen | 20.3 | 0.12% | 0.12% | 2.3 | 1.35-4.06 | 401.3 | 189-620 |
| Brunssum | 27.6 | 0.17% | 0.15% | 2.2 | 1.18-3.67 | 311.5 | 143-636 |
| Bunnik | 14.6 | 0.09% | 0.10% | 2.7 | 1.63-4.59 | 457.9 | 212-822 |
| Bunschoten | 21.3 | 0.13% | 0.14% | 2.6 | 1.65-4.15 | 437.7 | 160-748 |
| Buren | 26.0 | 0.16% | 0.18% | 2.8 | 1.76-4.30 | 448.4 | 196-877 |
| Capelle aan den IJssel | 66.4 | 0.40% | 0.45% | 2.8 | 2.08-4.21 | 524.0 | 222-888 |
| Castricum | 35.3 | 0.21% | 0.23% | 2.7 | 1.89-3.93 | 462.8 | 163-799 |
| Coevorden | 34.6 | 0.21% | 0.14% | 1.7 | 0.85-2.61 | 253.7 | 67-479 |
| Cranendonck | 19.9 | 0.12% | 0.12% | 2.5 | 1.36-5.03 | 409.9 | 172-801 |
| Cuijk | 24.3 | 0.15% | 0.16% | 2.7 | 1.70-4.33 | 477.5 | 218-916 |
| Culemborg | 27.9 | 0.17% | 0.19% | 2.7 | 1.80-3.92 | 497.7 | 231-824 |
| Dalfsen | 27.9 | 0.17% | 0.15% | 2.2 | 1.31-4.13 | 307.5 | 101-595 |
| Dantumadiel | 18.5 | 0.11% | 0.09% | 1.9 | 0.88-3.98 | 324.5 | 134-598 |
| De Bilt | 42.2 | 0.25% | 0.27% | 2.6 | 1.84-3.67 | 472.8 | 170-874 |
| De Fryske Marren | 51.0 | 0.31% | 0.29% | 2.3 | 1.47-3.46 | 340.8 | 117-674 |
| De Ronde Venen | 43.6 | 0.26% | 0.30% | 2.8 | 1.88-3.94 | 504.0 | 219-871 |
| De Wolden | 23.4 | 0.14% | 0.12% | 2.0 | 1.19-3.24 | 371.3 | 157-727 |
| Delft | 102.7 | 0.62% | 0.73% | 2.9 | 2.21-3.64 | 458.8 | 214-864 |
| Delfzijl | 24.2 | 0.15% | 0.08% | 1.4 | 0.72-3.33 | 241.7 | 74-515 |
| Den Helder | 55.1 | 0.33% | 0.31% | 2.3 | 1.51-3.29 | 390.5 | 154-713 |
| Deurne | 31.8 | 0.19% | 0.19% | 2.4 | 1.60-3.28 | 374.3 | 111-699 |
| Deventer | 99.6 | 0.60% | 0.52% | 2.1 | 1.47-3.05 | 329.9 | 108-668 |
| Diemen | 28.6 | 0.17% | 0.24% | 3.4 | 2.27-4.88 | 437.8 | 191-858 |
| Dinkelland | 25.9 | 0.16% | 0.11% | 1.8 | 0.88-3.71 | 328.9 | 108-655 |
| Doesburg | 10.5 | 0.06% | 0.06% | 2.4 | 1.17-4.66 | 387.2 | 146-691 |
| Doetinchem | 57.2 | 0.34% | 0.32% | 2.3 | 1.48-3.31 | 336.6 | 126-727 |
| Dongen | 25.5 | 0.15% | 0.16% | 2.6 | 1.60-4.38 | 449.6 | 178-781 |
| Dordrecht | 118.1 | 0.71% | 0.74% | 2.6 | 2.00-3.36 | 476.1 | 191-858 |
| Drechterland | 19.0 | 0.11% | 0.12% | 2.5 | 1.32-3.90 | 470.4 | 159-843 |
| Drimmelen | 26.5 | 0.16% | 0.18% | 2.7 | 1.76-4.55 | 437.2 | 183-734 |
| Dronten | 40.2 | 0.24% | 0.24% | 2.4 | 1.55-3.78 | 389.8 | 119-717 |
| Druten | 18.3 | 0.11% | 0.11% | 2.6 | 1.46-3.93 | 467.4 | 211-800 |
| Duiven | 24.9 | 0.15% | 0.15% | 2.4 | 1.40-3.94 | 431.3 | 165-759 |
| Echt-Susteren | 31.1 | 0.19% | 0.17% | 2.3 | 1.24-4.21 | 373.6 | 119-670 |
| Edam-Volendam | 35.7 | 0.21% | 0.23% | 2.6 | 1.60-4.24 | 403.7 | 166-743 |
| Ede | 115.2 | 0.69% | 0.66% | 2.3 | 1.73-3.00 | 456.5 | 179-829 |
| Eemnes | 8.6 | 0.05% | 0.05% | 2.4 | 1.24-4.28 | 500.7 | 236-823 |
| Eersel | 18.6 | 0.11% | 0.12% | 2.7 | 1.65-3.91 | 466.6 | 199-940 |
| Eijsden-Margraten | 25.1 | 0.15% | 0.14% | 2.3 | 1.20-3.76 | 318.5 | 106-621 |

Continued on next page

| Municipality | Population size (×1000) | Population percentage | Percentage of transmissions | Transmission risk | (90% range) | Seed Risk | (90% range) |
|---|---|---|---|---|---|---|---|
| Eindhoven | 230.5 | 1.38% | 1.41% | 2.5 | 1.86-3.23 | 402.0 | 149-760 |
| Elburg | 22.5 | 0.14% | 0.15% | 2.7 | 1.57-4.76 | 404.6 | 148-780 |
| Emmen | 106.4 | 0.64% | 0.44% | 1.7 | 1.04-2.76 | 250.8 | 46-575 |
| Enkhuizen | 18.0 | 0.11% | 0.12% | 2.7 | 1.48-4.44 | 463.5 | 195-812 |
| Enschede | 158.2 | 0.95% | 0.64% | 1.7 | 1.20-2.15 | 268.1 | 80-650 |
| Epe | 32.8 | 0.20% | 0.20% | 2.4 | 1.50-3.94 | 338.8 | 134-691 |
| Ermelo | 26.3 | 0.16% | 0.18% | 2.8 | 1.68-4.35 | 466.4 | 229-806 |
| Etten-Leur | 43.2 | 0.26% | 0.28% | 2.7 | 1.89-3.84 | 419.0 | 156-774 |
| Geertruidenberg | 21.0 | 0.13% | 0.14% | 2.7 | 1.73-3.91 | 475.9 | 207-849 |
| Geldrop-Mierlo | 39.1 | 0.23% | 0.25% | 2.6 | 1.63-3.88 | 498.2 | 170-928 |
| Gemert-Bakel | 30.0 | 0.18% | 0.19% | 2.7 | 1.46-4.64 | 336.4 | 111-627 |
| Gennep | 16.7 | 0.10% | 0.09% | 2.3 | 1.26-4.31 | 430.4 | 204-855 |
| Gilze en Rijen | 25.9 | 0.16% | 0.17% | 2.6 | 1.52-4.62 | 393.1 | 155-822 |
| Goeree-Overflakkee | 49.2 | 0.30% | 0.26% | 2.2 | 1.53-3.49 | 384.9 | 149-729 |
| Goes | 37.1 | 0.22% | 0.18% | 2.0 | 1.04-2.98 | 306.2 | 108-668 |
| Goirle | 23.1 | 0.14% | 0.14% | 2.6 | 1.62-4.14 | 484.2 | 216-808 |
| Gooise Meren | 57.1 | 0.34% | 0.39% | 2.8 | 2.08-4.33 | 496.9 | 228-841 |
| Gorinchem | 35.9 | 0.22% | 0.26% | 3.0 | 1.99-4.65 | 485.3 | 202-833 |
| Gouda | 72.6 | 0.44% | 0.49% | 2.8 | 2.03-4.03 | 427.6 | 194-707 |
| Grave | 12.1 | 0.07% | 0.08% | 2.6 | 1.37-4.66 | 425.1 | 217-756 |
| Groningen | 203.2 | 1.22% | 0.94% | 1.9 | 1.37-2.77 | 299.2 | 83-557 |
| Gulpen-Wittem | 13.8 | 0.08% | 0.07% | 2.1 | 1.01-4.20 | 352.4 | 179-576 |
| Haaksbergen | 23.6 | 0.14% | 0.11% | 1.9 | 1.05-3.21 | 330.3 | 116-599 |
| Haaren | 13.6 | 0.08% | 0.09% | 2.8 | 1.54-4.44 | 422.5 | 189-727 |
| Haarlem | 160.5 | 0.96% | 1.02% | 2.6 | 2.11-3.12 | 480.3 | 219-806 |
| Haarlemmermeer | 147.3 | 0.88% | 0.99% | 2.8 | 2.08-3.76 | 501.3 | 265-865 |
| Halderberge | 29.8 | 0.18% | 0.16% | 2.2 | 1.45-3.47 | 402.4 | 127-902 |
| Hardenberg | 60.3 | 0.36% | 0.31% | 2.1 | 1.42-3.25 | 344.3 | 99-743 |
| Harderwijk | 46.7 | 0.28% | 0.28% | 2.5 | 1.69-3.72 | 399.6 | 166-690 |
| Hardinxveld-Giessendam | 17.5 | 0.11% | 0.11% | 2.6 | 1.67-3.62 | 480.2 | 179-941 |
| Harlingen | 15.3 | 0.09% | 0.07% | 2.0 | 1.00-3.73 | 337.7 | 135-652 |
| Hattem | 11.6 | 0.07% | 0.07% | 2.4 | 1.32-4.11 | 394.4 | 171-721 |
| Heemskerk | 38.8 | 0.23% | 0.30% | 3.1 | 2.17-4.14 | 530.4 | 210-904 |
| Heemstede | 26.8 | 0.16% | 0.19% | 3.0 | 2.01-4.86 | 522.5 | 218-984 |
| Heerde | 18.1 | 0.11% | 0.10% | 2.3 | 1.36-3.75 | 377.5 | 194-650 |
| Heerenveen | 49.6 | 0.30% | 0.26% | 2.1 | 1.41-3.42 | 328.1 | 114-654 |
| Heerhugowaard | 56.0 | 0.34% | 0.33% | 2.4 | 1.68-3.45 | 406.4 | 150-837 |
| Heerlen | 86.6 | 0.52% | 0.43% | 2.0 | 1.46-2.66 | 345.3 | 83-680 |
| Heeze-Leende | 15.5 | 0.09% | 0.09% | 2.4 | 1.25-4.21 | 400.5 | 179-686 |
| Heiloo | 23.0 | 0.14% | 0.14% | 2.6 | 1.72-3.87 | 451.6 | 180-734 |
| Hellendoorn | 35.4 | 0.21% | 0.20% | 2.3 | 1.30-3.94 | 309.9 | 143-549 |
| Hellevoetsluis | 39.6 | 0.24% | 0.26% | 2.7 | 1.53-4.80 | 394.2 | 163-691 |
| Helmond | 90.9 | 0.55% | 0.52% | 2.3 | 1.66-3.30 | 368.4 | 128-796 |
| Hendrik-Ido-Ambacht | 30.4 | 0.18% | 0.19% | 2.6 | 1.83-3.88 | 454.1 | 189-869 |
| Hengelo | 80.3 | 0.48% | 0.41% | 2.1 | 1.37-3.17 | 266.2 | 69-591 |
| Het Hogeland | 11.2 | 0.07% | 0.06% | 2.1 | 0.63-5.01 | 269.1 | 0-668 |
| Heumen | 16.0 | 0.10% | 0.09% | 2.4 | 1.43-3.40 | 418.8 | 155-747 |
| Heusden | 43.6 | 0.26% | 0.30% | 2.8 | 1.81-3.84 | 478.9 | 231-883 |
| Hillegom | 21.4 | 0.13% | 0.14% | 2.8 | 1.77-4.12 | 514.9 | 236-888 |
| Hilvarenbeek | 14.9 | 0.09% | 0.13% | 3.7 | 2.19-5.88 | 425.9 | 150-833 |
| Hilversum | 89.7 | 0.54% | 0.62% | 2.8 | 2.07-3.92 | 480.8 | 219-974 |
| Hoeksche Waard | 11.2 | 0.07% | 0.07% | 2.4 | 1.14-4.10 | 445.9 | 188-895 |
| Hof van Twente | 34.5 | 0.21% | 0.17% | 2.0 | 1.37-3.05 | 300.2 | 129-590 |
| Hollands Kroon | 46.9 | 0.28% | 0.26% | 2.3 | 1.37-4.00 | 331.0 | 103-628 |
| Hoogeveen | 55.0 | 0.33% | 0.28% | 2.0 | 1.22-3.29 | 293.9 | 100-572 |
| Hoorn | 72.6 | 0.44% | 0.46% | 2.6 | 1.92-3.85 | 409.9 | 176-723 |
| Horst aan de Maas | 41.7 | 0.25% | 0.23% | 2.3 | 1.53-3.32 | 378.0 | 155-688 |
| Houten | 49.5 | 0.30% | 0.34% | 2.8 | 1.91-3.83 | 469.4 | 181-776 |
| Huizen | 40.9 | 0.25% | 0.28% | 2.8 | 1.88-4.15 | 469.5 | 227-915 |
| Hulst | 27.0 | 0.16% | 0.15% | 2.2 | 1.25-4.04 | 341.9 | 101-606 |
| IJsselstein | 33.6 | 0.20% | 0.26% | 3.1 | 2.15-4.71 | 448.5 | 200-746 |
| Kaag en Braassem | 26.4 | 0.16% | 0.19% | 2.9 | 1.94-4.31 | 437.0 | 163-772 |
| Kampen | 53.2 | 0.32% | 0.29% | 2.2 | 1.58-3.13 | 395.0 | 126-767 |

Continued on next page

| Municipality | Population size (×1000) | Population percentage | Percentage of transmissions | Transmission risk | (90% range) | Seed Risk | (90% range) |
|---|---|---|---|---|---|---|---|
| Kapelle | 12.4 | 0.07% | 0.06% | 1.8 | 0.97-3.19 | 304.5 | 117-607 |
| Katwijk | 64.6 | 0.39% | 0.47% | 3.0 | 2.10-4.24 | 492.5 | 247-816 |
| Kerkrade | 45.1 | 0.27% | 0.22% | 2.0 | 1.23-3.15 | 349.0 | 101-689 |
| Koggenland | 22.4 | 0.13% | 0.13% | 2.4 | 1.49-3.47 | 418.4 | 181-824 |
| Krimpen aan den IJssel | 28.8 | 0.17% | 0.22% | 3.1 | 2.31-4.28 | 606.9 | 300-1,031 |
| Krimpenerwaard | 55.5 | 0.33% | 0.35% | 2.6 | 1.77-3.36 | 476.1 | 220-797 |
| Laarbeek | 21.8 | 0.13% | 0.15% | 2.9 | 1.71-5.19 | 434.7 | 166-822 |
| Landerd | 14.8 | 0.09% | 0.09% | 2.5 | 1.42-4.36 | 448.4 | 165-743 |
| Landgraaf | 37.0 | 0.22% | 0.18% | 1.9 | 1.20-3.26 | 327.6 | 101-593 |
| Landsmeer | 11.0 | 0.07% | 0.08% | 3.0 | 1.56-4.50 | 504.5 | 208-899 |
| Langedijk | 27.5 | 0.17% | 0.16% | 2.3 | 1.36-4.12 | 382.2 | 155-691 |
| Lansingerland | 61.1 | 0.37% | 0.43% | 2.9 | 2.12-4.18 | 506.9 | 231-835 |
| Laren | 10.7 | 0.06% | 0.07% | 2.6 | 1.36-4.54 | 503.7 | 234-908 |
| Leeuwarden | 122.4 | 0.73% | 0.60% | 2.0 | 1.39-2.87 | 308.5 | 122-672 |
| Leiden | 124.1 | 0.75% | 0.84% | 2.8 | 2.09-3.53 | 499.4 | 196-876 |
| Leiderdorp | 26.7 | 0.16% | 0.18% | 2.8 | 1.81-3.95 | 490.9 | 217-882 |
| Leidschendam-Voorburg | 74.8 | 0.45% | 0.50% | 2.8 | 2.14-3.85 | 516.8 | 266-926 |
| Lelystad | 77.6 | 0.47% | 0.46% | 2.4 | 1.61-3.43 | 393.6 | 137-809 |
| Leudal | 35.3 | 0.21% | 0.19% | 2.1 | 1.43-3.47 | 345.3 | 110-652 |
| Leusden | 29.2 | 0.18% | 0.20% | 2.8 | 1.70-5.20 | 543.9 | 264-1,015 |
| Lingewaard | 45.9 | 0.28% | 0.29% | 2.6 | 1.79-3.78 | 407.7 | 159-741 |
| Lisse | 22.4 | 0.13% | 0.16% | 2.8 | 1.94-4.30 | 495.1 | 218-934 |
| Lochem | 33.1 | 0.20% | 0.18% | 2.2 | 1.35-3.74 | 326.2 | 140-631 |
| Loon op Zand | 22.6 | 0.14% | 0.15% | 2.8 | 1.92-4.08 | 498.7 | 242-892 |
| Lopik | 13.9 | 0.08% | 0.10% | 2.9 | 1.70-5.10 | 423.7 | 173-821 |
| Loppersum | 9.0 | 0.05% | 0.03% | 1.5 | 0.42-3.58 | 301.7 | 83-681 |
| Losser | 22.2 | 0.13% | 0.09% | 1.6 | 0.70-3.04 | 259.8 | 48-645 |
| Maasdriel | 24.0 | 0.14% | 0.17% | 2.8 | 1.70-4.93 | 416.4 | 192-724 |
| Maasgouw | 23.2 | 0.14% | 0.13% | 2.3 | 1.21-3.86 | 312.3 | 119-547 |
| Maassluis | 32.2 | 0.19% | 0.23% | 2.9 | 1.92-3.86 | 441.2 | 170-788 |
| Maastricht | 121.7 | 0.73% | 0.58% | 1.9 | 1.38-2.78 | 298.2 | 106-579 |
| Medemblik | 44.2 | 0.27% | 0.24% | 2.2 | 1.51-3.45 | 401.1 | 134-747 |
| Meerssen | 18.5 | 0.11% | 0.10% | 2.2 | 1.16-3.70 | 379.5 | 114-740 |
| Meierijstad | 80.3 | 0.48% | 0.51% | 2.6 | 1.98-3.60 | 459.7 | 216-776 |
| Meppel | 33.0 | 0.20% | 0.17% | 2.1 | 1.33-3.26 | 351.1 | 140-692 |
| Middelburg | 48.3 | 0.29% | 0.19% | 1.6 | 0.97-2.47 | 275.3 | 62-600 |
| Midden-Delfland | 19.1 | 0.11% | 0.17% | 3.6 | 2.22-6.53 | 470.6 | 185-744 |
| Midden-Drenthe | 32.5 | 0.20% | 0.16% | 2.1 | 1.04-3.35 | 275.1 | 71-552 |
| Midden-Groningen | 60.5 | 0.36% | 0.26% | 1.7 | 1.09-3.16 | 293.3 | 51-612 |
| Mill en Sint Hubert | 10.5 | 0.06% | 0.07% | 2.7 | 1.45-4.97 | 421.4 | 175-728 |
| Moerdijk | 36.6 | 0.22% | 0.23% | 2.6 | 1.90-3.93 | 448.0 | 183-789 |
| Molenlanden | 11.2 | 0.07% | 0.07% | 2.7 | 1.57-3.70 | 518.7 | 177-1,066 |
| Montferland | 35.3 | 0.21% | 0.19% | 2.2 | 1.34-3.16 | 335.6 | 94-615 |
| Montfoort | 13.4 | 0.08% | 0.09% | 2.8 | 1.55-4.44 | 472.9 | 221-851 |
| Mook en Middelaar | 7.3 | 0.04% | 0.04% | 2.2 | 1.20-3.58 | 411.0 | 185-736 |
| Neder-Betuwe | 23.3 | 0.14% | 0.14% | 2.4 | 1.56-3.75 | 465.8 | 148-877 |
| Nederweert | 16.6 | 0.10% | 0.10% | 2.5 | 1.50-4.18 | 395.9 | 146-704 |
| Nieuwegein | 62.5 | 0.38% | 0.45% | 2.9 | 2.27-4.05 | 537.1 | 265-904 |
| Nieuwkoop | 28.0 | 0.17% | 0.19% | 2.8 | 1.88-4.18 | 465.2 | 184-834 |
| Nijkerk | 42.3 | 0.25% | 0.27% | 2.6 | 1.68-3.77 | 426.1 | 200-756 |
| Nijmegen | 176.6 | 1.06% | 1.04% | 2.4 | 1.81-3.46 | 403.0 | 142-714 |
| Nissewaard | 84.2 | 0.51% | 0.52% | 2.5 | 1.78-3.50 | 403.8 | 200-712 |
| Noardeast-FryslÃ¢n | 11.2 | 0.07% | 0.04% | 1.5 | 0.57-3.05 | 268.7 | 54-530 |
| Noord-Beveland | 6.7 | 0.04% | 0.03% | 1.7 | 0.57-3.45 | 292.6 | 70-643 |
| Noordenveld | 30.8 | 0.18% | 0.14% | 1.8 | 1.14-3.30 | 289.5 | 101-566 |
| Noordoostpolder | 46.3 | 0.28% | 0.25% | 2.2 | 1.58-3.81 | 315.1 | 103-621 |
| Noordwijk | 25.8 | 0.15% | 0.18% | 2.8 | 1.63-4.54 | 466.1 | 203-802 |
| Nuenen, Gerwen en Nederwetten | 22.7 | 0.14% | 0.14% | 2.4 | 1.55-4.17 | 418.3 | 139-790 |
| Nunspeet | 27.1 | 0.16% | 0.17% | 2.5 | 1.53-4.26 | 421.7 | 180-736 |

Continued on next page

| Municipality | Population size (×1000) | Population percentage | Percentage of transmissions | Transmission risk | (90% range) | Seed Risk | (90% range) |
|---|---|---|---|---|---|---|---|
| Oegstgeest | 23.9 | 0.14% | 0.20% | 3.5 | 2.25-4.92 | 491.4 | 226-854 |
| Oirschot | 18.1 | 0.11% | 0.11% | 2.6 | 1.44-3.93 | 480.9 | 202-907 |
| Oisterwijk | 25.4 | 0.15% | 0.16% | 2.7 | 1.42-5.27 | 413.7 | 126-728 |
| Oldambt | 37.5 | 0.23% | 0.16% | 1.8 | 0.89-3.66 | 196.5 | 42-447 |
| Oldebroek | 23.1 | 0.14% | 0.14% | 2.5 | 1.38-4.33 | 379.9 | 129-739 |
| Oldenzaal | 31.1 | 0.19% | 0.13% | 1.8 | 1.05-2.83 | 298.9 | 93-573 |
| Olst-Wijhe | 17.6 | 0.11% | 0.10% | 2.3 | 1.26-3.71 | 413.9 | 132-771 |
| Ommen | 17.4 | 0.10% | 0.10% | 2.3 | 1.01-4.86 | 357.8 | 107-674 |
| Oost Gelre | 29.2 | 0.18% | 0.14% | 2.0 | 1.25-3.10 | 301.4 | 92-677 |
| Oosterhout | 54.7 | 0.33% | 0.36% | 2.7 | 1.91-3.92 | 486.4 | 192-895 |
| Ooststellingwerf | 24.9 | 0.15% | 0.12% | 2.0 | 1.11-3.16 | 307.7 | 123-599 |
| Oostzaan | 9.2 | 0.06% | 0.07% | 3.1 | 1.31-4.78 | 501.7 | 254-877 |
| Opmeer | 11.2 | 0.07% | 0.07% | 2.6 | 1.27-5.07 | 472.8 | 210-784 |
| Opsterland | 29.4 | 0.18% | 0.15% | 2.0 | 1.16-3.20 | 324.8 | 108-631 |
| Oss | 91.0 | 0.55% | 0.56% | 2.5 | 1.80-3.40 | 454.9 | 182-934 |
| Oude IJsselstreek | 38.8 | 0.23% | 0.20% | 2.1 | 1.24-3.47 | 326.9 | 79-608 |
| Ouder-Amstel | 13.3 | 0.08% | 0.10% | 3.2 | 1.87-5.26 | 540.1 | 219-909 |
| Oudewater | 9.7 | 0.06% | 0.07% | 2.9 | 1.59-4.91 | 522.5 | 262-877 |
| Overbetuwe | 47.0 | 0.28% | 0.28% | 2.4 | 1.74-3.18 | 450.1 | 168-863 |
| Papendrecht | 31.8 | 0.19% | 0.20% | 2.5 | 1.73-3.59 | 482.7 | 203-882 |
| Peel en Maas | 42.9 | 0.26% | 0.24% | 2.3 | 1.39-3.88 | 360.7 | 73-639 |
| Pekela | 11.9 | 0.07% | 0.05% | 1.8 | 0.69-3.43 | 248.4 | 78-516 |
| Pijnacker-Nootdorp | 53.6 | 0.32% | 0.45% | 3.4 | 2.60-4.76 | 540.1 | 196-986 |
| Purmerend | 79.6 | 0.48% | 0.53% | 2.7 | 1.89-4.05 | 447.5 | 162-826 |
| Putten | 23.8 | 0.14% | 0.16% | 2.7 | 1.76-4.08 | 480.9 | 188-886 |
| Raalte | 36.9 | 0.22% | 0.19% | 2.1 | 1.32-3.50 | 372.3 | 126-753 |
| Reimerswaal | 22.2 | 0.13% | 0.11% | 2.0 | 1.09-3.70 | 332.2 | 124-722 |
| Renkum | 30.9 | 0.19% | 0.21% | 2.7 | 1.72-4.33 | 467.1 | 166-893 |
| Renswoude | 4.6 | 0.03% | 0.04% | 3.6 | 1.07-8.60 | 487.7 | 225-795 |
| Reusel-De Mierden | 12.6 | 0.08% | 0.08% | 2.4 | 1.30-4.23 | 410.9 | 161-728 |
| Rheden | 43.3 | 0.26% | 0.24% | 2.3 | 1.55-3.56 | 337.7 | 110-619 |
| Rhenen | 19.4 | 0.12% | 0.14% | 2.9 | 1.86-4.92 | 483.9 | 230-928 |
| Ridderkerk | 45.7 | 0.27% | 0.29% | 2.6 | 1.82-3.82 | 433.7 | 198-816 |
| Rijssen-Holten | 37.8 | 0.23% | 0.20% | 2.2 | 1.29-3.72 | 319.8 | 78-630 |
| Rijswijk | 52.7 | 0.32% | 0.38% | 3.0 | 2.20-4.03 | 469.5 | 254-765 |
| Roerdalen | 20.1 | 0.12% | 0.10% | 2.0 | 1.29-3.70 | 387.1 | 139-736 |
| Roermond | 57.4 | 0.34% | 0.31% | 2.2 | 1.37-3.36 | 376.1 | 150-749 |
| Roosendaal | 76.6 | 0.46% | 0.44% | 2.4 | 1.70-3.36 | 402.2 | 133-751 |
| Rotterdam | 641.3 | 3.85% | 4.16% | 2.7 | 2.34-3.06 | 466.1 | 226-798 |
| Rozendaal | 1.3 | 0.01% | 0.00% | 1.3 | 0.15-2.91 | 426.2 | 174-671 |
| Rucphen | 22.2 | 0.13% | 0.15% | 2.7 | 1.49-4.67 | 428.5 | 158-809 |
| Schagen | 45.9 | 0.28% | 0.30% | 2.7 | 1.69-5.55 | 488.8 | 179-880 |
| Scherpenzeel | 9.5 | 0.06% | 0.06% | 2.5 | 1.41-4.71 | 413.0 | 182-765 |
| Schiedam | 77.4 | 0.46% | 0.63% | 3.3 | 2.60-4.26 | 469.8 | 176-808 |
| Schiermonnikoog | 0.5 | 0.00% | 0.00% | 0.9 | 0.00-2.48 | 410.2 | 156-765 |
| Schouwen-Duiveland | 33.3 | 0.20% | 0.16% | 1.9 | 1.15-2.86 | 346.7 | 102-759 |
| Simpelveld | 9.9 | 0.06% | 0.05% | 2.1 | 0.85-4.23 | 331.9 | 105-666 |
| Sint Anthonis | 11.2 | 0.07% | 0.07% | 2.5 | 1.33-4.12 | 429.4 | 145-832 |
| Sint-Michielsgestel | 28.5 | 0.17% | 0.20% | 2.9 | 1.87-4.90 | 464.1 | 196-789 |
| Sittard-Geleen | 92.2 | 0.55% | 0.47% | 2.1 | 1.44-3.08 | 329.3 | 85-671 |
| Sliedrecht | 24.6 | 0.15% | 0.18% | 2.9 | 1.85-4.71 | 475.4 | 235-902 |
| Sluis | 23.0 | 0.14% | 0.09% | 1.6 | 0.77-2.57 | 295.4 | 56-643 |
| Smallingerland | 55.4 | 0.33% | 0.29% | 2.1 | 1.40-3.60 | 303.1 | 93-583 |
| Soest | 45.6 | 0.27% | 0.29% | 2.6 | 1.86-3.73 | 501.6 | 211-890 |
| Someren | 18.8 | 0.11% | 0.12% | 2.6 | 1.36-4.43 | 341.9 | 121-592 |
| Son en Breugel | 16.2 | 0.10% | 0.11% | 2.7 | 1.64-3.89 | 440.9 | 218-770 |
| Stadskanaal | 31.4 | 0.19% | 0.14% | 1.8 | 0.87-3.04 | 256.7 | 51-604 |
| Staphorst | 16.4 | 0.10% | 0.08% | 2.0 | 1.03-3.21 | 297.2 | 119-624 |
| Stede Broec | 21.3 | 0.13% | 0.12% | 2.4 | 1.47-3.77 | 366.3 | 137-689 |
| Steenbergen | 25.0 | 0.15% | 0.15% | 2.5 | 1.40-4.33 | 302.1 | 105-513 |
| Steenwijkerland | 43.3 | 0.26% | 0.24% | 2.2 | 1.51-3.30 | 365.3 | 168-684 |
| Stein | 24.4 | 0.15% | 0.13% | 2.3 | 1.35-4.02 | 365.9 | 137-637 |
| Stichtse Vecht | 63.9 | 0.38% | 0.42% | 2.7 | 1.91-4.04 | 442.8 | 195-736 |

Continued on next page

| Municipality | Population size (×1000) | Population percentage | Percentage of transmissions | Transmission risk | (90% range) | Seed Risk | (90% range) |
|---|---|---|---|---|---|---|---|
| SÃºdwest-FryslÃ¢n | 89.0 | 0.53% | 0.44% | 2.0 | 1.40-2.78 | 340.7 | 90-682 |
| Terneuzen | 54.1 | 0.32% | 0.25% | 1.9 | 1.03-3.00 | 313.0 | 94-663 |
| Terschelling | 4.5 | 0.03% | 0.02% | 2.1 | 0.48-4.68 | 299.1 | 100-595 |
| Texel | 13.0 | 0.08% | 0.07% | 2.1 | 0.87-3.39 | 358.3 | 112-754 |
| Teylingen | 36.5 | 0.22% | 0.24% | 2.7 | 1.68-4.20 | 499.3 | 234-794 |
| Tholen | 25.2 | 0.15% | 0.15% | 2.4 | 1.22-4.32 | 367.8 | 108-784 |
| Tiel | 41.5 | 0.25% | 0.26% | 2.5 | 1.84-3.53 | 467.1 | 208-905 |
| Tilburg | 216.5 | 1.30% | 1.24% | 2.3 | 1.83-3.24 | 458.0 | 193-816 |
| Tubbergen | 20.6 | 0.12% | 0.10% | 2.0 | 1.03-3.29 | 308.1 | 104-637 |
| Twenterand | 33.6 | 0.20% | 0.16% | 2.0 | 1.18-3.47 | 318.9 | 92-575 |
| Tynaarlo | 33.2 | 0.20% | 0.13% | 1.6 | 0.90-2.71 | 236.4 | 65-489 |
| Tytsjerksteradiel | 31.3 | 0.19% | 0.15% | 2.0 | 1.07-3.31 | 332.8 | 118-617 |
| Uden | 41.2 | 0.25% | 0.26% | 2.6 | 1.72-3.88 | 390.9 | 106-685 |
| Uitgeest | 13.0 | 0.08% | 0.09% | 2.8 | 1.59-4.44 | 441.0 | 157-852 |
| Uithoorn | 28.9 | 0.17% | 0.20% | 2.9 | 1.78-4.53 | 478.1 | 206-853 |
| Urk | 20.2 | 0.12% | 0.13% | 2.6 | 1.34-4.30 | 364.0 | 112-729 |
| Utrecht | 350.4 | 2.10% | 2.19% | 2.6 | 2.06-3.11 | 440.2 | 212-792 |
| Utrechtse Heuvelrug | 49.0 | 0.29% | 0.30% | 2.5 | 1.72-3.67 | 410.9 | 134-785 |
| Vaals | 9.6 | 0.06% | 0.05% | 2.0 | 0.90-3.84 | 360.3 | 126-720 |
| Valkenburg aan de Geul | 16.0 | 0.10% | 0.08% | 1.9 | 0.95-3.21 | 360.7 | 127-692 |
| Valkenswaard | 30.4 | 0.18% | 0.19% | 2.5 | 1.67-4.09 | 421.4 | 175-721 |
| Veendam | 26.9 | 0.16% | 0.12% | 1.8 | 0.84-3.04 | 279.6 | 55-637 |
| Veenendaal | 64.9 | 0.39% | 0.40% | 2.5 | 1.78-3.60 | 465.0 | 209-970 |
| Veere | 21.2 | 0.13% | 0.09% | 1.8 | 0.78-3.75 | 250.6 | 66-541 |
| Veldhoven | 44.6 | 0.27% | 0.26% | 2.4 | 1.62-3.40 | 365.8 | 171-713 |
| Velsen | 67.7 | 0.41% | 0.45% | 2.7 | 2.03-3.97 | 463.4 | 170-845 |
| Venlo | 101.0 | 0.61% | 0.53% | 2.1 | 1.49-2.98 | 314.7 | 97-578 |
| Venray | 42.8 | 0.26% | 0.23% | 2.2 | 1.42-3.06 | 347.1 | 69-676 |
| Vijfheerenlanden | 11.2 | 0.07% | 0.08% | 2.9 | 1.59-4.65 | 532.3 | 210-976 |
| Vlaardingen | 71.8 | 0.43% | 0.55% | 3.1 | 2.35-4.54 | 404.4 | 140-716 |
| Vlieland | 0.6 | 0.00% | 0.00% | 1.0 | 0.00-3.06 | 406.0 | 145-759 |
| Vlissingen | 44.1 | 0.26% | 0.19% | 1.7 | 1.04-2.84 | 229.7 | 66-529 |
| Voerendaal | 12.0 | 0.07% | 0.06% | 2.1 | 1.03-4.38 | 372.9 | 117-709 |
| Voorschoten | 25.0 | 0.15% | 0.17% | 2.8 | 1.85-4.52 | 461.7 | 225-810 |
| Voorst | 23.9 | 0.14% | 0.13% | 2.3 | 1.41-3.23 | 363.6 | 148-666 |
| Vught | 26.0 | 0.16% | 0.17% | 2.7 | 1.83-4.18 | 477.1 | 230-850 |
| Waadhoeke | 45.6 | 0.27% | 0.24% | 2.1 | 1.32-3.16 | 323.6 | 97-595 |
| Waalre | 16.9 | 0.10% | 0.12% | 2.8 | 1.56-4.67 | 472.5 | 230-902 |
| Waalwijk | 47.7 | 0.29% | 0.29% | 2.5 | 1.74-3.50 | 423.4 | 136-786 |
| Waddinxveen | 27.9 | 0.17% | 0.20% | 3.0 | 2.05-4.42 | 477.1 | 224-865 |
| Wageningen | 38.1 | 0.23% | 0.25% | 2.7 | 1.96-3.47 | 429.3 | 152-771 |
| Wassenaar | 25.6 | 0.15% | 0.22% | 3.5 | 2.24-5.60 | 418.8 | 188-691 |
| Waterland | 16.8 | 0.10% | 0.12% | 2.9 | 1.87-4.33 | 464.3 | 142-802 |
| Weert | 49.1 | 0.29% | 0.24% | 2.0 | 1.46-3.05 | 309.3 | 119-533 |
| Weesp | 18.8 | 0.11% | 0.14% | 3.0 | 1.78-4.35 | 453.4 | 222-804 |
| West Betuwe | 11.2 | 0.07% | 0.08% | 2.8 | 1.39-5.56 | 478.8 | 147-857 |
| West Maas en Waal | 18.5 | 0.11% | 0.13% | 2.8 | 1.50-4.80 | 418.3 | 168-710 |
| Westerkwartier | 11.2 | 0.07% | 0.05% | 1.9 | 0.60-4.25 | 218.9 | 77-388 |
| Westerveld | 18.9 | 0.11% | 0.08% | 1.8 | 0.99-3.00 | 296.1 | 94-544 |
| Westervoort | 14.5 | 0.09% | 0.08% | 2.4 | 1.31-3.93 | 405.5 | 193-743 |
| Westerwolde | 24.8 | 0.15% | 0.10% | 1.6 | 0.76-3.69 | 224.0 | 44-533 |
| Westland | 108.3 | 0.65% | 0.65% | 2.5 | 1.93-3.09 | 462.1 | 207-914 |
| Weststellingwerf | 25.4 | 0.15% | 0.13% | 2.2 | 1.26-4.14 | 345.0 | 104-660 |
| Westvoorne | 14.3 | 0.09% | 0.08% | 2.2 | 1.10-4.09 | 334.3 | 110-655 |
| Wierden | 23.9 | 0.14% | 0.14% | 2.4 | 1.06-4.82 | 331.3 | 134-605 |
| Wijchen | 40.4 | 0.24% | 0.26% | 2.7 | 1.84-3.74 | 472.0 | 214-842 |
| Wijdemeren | 23.4 | 0.14% | 0.17% | 2.9 | 1.80-4.76 | 494.0 | 205-803 |
| Wijk bij Duurstede | 23.3 | 0.14% | 0.16% | 2.8 | 1.73-4.50 | 463.2 | 194-829 |
| Winterswijk | 28.5 | 0.17% | 0.14% | 2.1 | 1.13-3.74 | 349.5 | 94-689 |
| Woensdrecht | 21.4 | 0.13% | 0.12% | 2.3 | 1.30-4.15 | 410.8 | 127-700 |
| Woerden | 51.5 | 0.31% | 0.34% | 2.7 | 1.78-3.60 | 429.4 | 188-735 |

Continued on next page

| Municipality | Population size (×1000) | Population percentage | Percentage of transmissions | Transmission risk | (90% range) | Seed Risk | (90% range) |
|---|---|---|---|---|---|---|---|
| Wormerland | 15.8 | 0.09% | 0.13% | 3.3 | 1.68-5.57 | 538.6 | 305-888 |
| Woudenberg | 12.6 | 0.08% | 0.10% | 3.2 | 1.72-7.91 | 459.5 | 175-836 |
| Zaanstad | 155.1 | 0.93% | 1.01% | 2.7 | 2.08-3.46 | 467.1 | 192-786 |
| Zaltbommel | 27.9 | 0.17% | 0.19% | 2.7 | 1.87-3.97 | 475.2 | 180-863 |
| Zandvoort | 16.3 | 0.10% | 0.11% | 2.7 | 1.61-4.75 | 433.0 | 197-732 |
| Zeewolde | 21.8 | 0.13% | 0.15% | 2.7 | 1.60-4.51 | 443.2 | 202-791 |
| Zeist | 63.6 | 0.38% | 0.39% | 2.5 | 1.83-3.06 | 438.6 | 199-821 |
| Zevenaar | 43.0 | 0.26% | 0.23% | 2.2 | 1.47-2.82 | 379.6 | 92-731 |
| Zoetermeer | 124.4 | 0.75% | 0.84% | 2.8 | 2.16-3.59 | 523.6 | 242-872 |
| Zoeterwoude | 8.1 | 0.05% | 0.06% | 2.8 | 1.41-5.10 | 500.6 | 228-865 |
| Zuidplas | 42.1 | 0.25% | 0.30% | 2.9 | 2.20-4.40 | 506.5 | 247-951 |
| Zundert | 21.4 | 0.13% | 0.13% | 2.6 | 1.61-4.22 | 466.3 | 240-830 |
| Zutphen | 47.0 | 0.28% | 0.25% | 2.2 | 1.51-3.01 | 362.9 | 120-671 |
| Zwartewaterland | 22.0 | 0.13% | 0.13% | 2.4 | 1.36-3.58 | 364.4 | 123-674 |
| Zwijndrecht | 44.3 | 0.27% | 0.30% | 2.7 | 1.94-3.92 | 453.5 | 216-802 |
| Zwolle | 126.6 | 0.76% | 0.65% | 2.1 | 1.52-3.15 | 342.0 | 108-627 |

**Supplementary Table SI.2:** The full list of population (data from 2019) and 17-days transmission risk scores and seed risk scores of the municipalities in the Netherlands after an initial introduction in students.

| Municipality | Population size (×1000) | Population percentage | Percentage of transmissions | Transmission risk | (90% range) | Seed Risk | (90% range) |
|---|---|---|---|---|---|---|---|
| ’s-Gravenhage | 535.6 | 3.22% | 3.48% | 6.6 | 5.70-7.57 | 1,080.5 | 525-2,123 |
| ’s-Hertogenbosch | 153.5 | 0.92% | 0.94% | 6.2 | 5.16-7.66 | 1,060.8 | 495-1,779 |
| Aa en Hunze | 24.8 | 0.15% | 0.13% | 5.1 | 2.95-9.96 | 632.2 | 190-1,171 |
| Aalsmeer | 31.0 | 0.19% | 0.21% | 6.9 | 5.29-8.93 | 1,281.9 | 592-2,648 |
| Aalten | 26.5 | 0.16% | 0.13% | 4.8 | 3.09-7.84 | 639.2 | 170-1,501 |
| Achtkarspelen | 27.5 | 0.17% | 0.13% | 4.9 | 3.24-8.01 | 635.5 | 209-1,275 |
| Alblasserdam | 19.7 | 0.12% | 0.14% | 7.1 | 5.03-9.89 | 1,222.4 | 564-2,229 |
| Albrandswaard | 24.8 | 0.15% | 0.17% | 6.9 | 5.01-9.98 | 1,134.9 | 510-2,065 |
| Alkmaar | 108.2 | 0.65% | 0.61% | 5.7 | 4.62-7.04 | 1,071.5 | 413-2,002 |
| Almelo | 72.4 | 0.43% | 0.35% | 4.9 | 3.66-6.63 | 778.1 | 233-1,645 |
| Almere | 206.2 | 1.24% | 1.29% | 6.3 | 5.38-7.43 | 1,139.9 | 527-1,983 |
| Alphen aan den Rijn | 110.2 | 0.66% | 0.74% | 6.8 | 5.64-8.22 | 1,124.7 | 681-1,791 |
| Alphen-Chaam | 9.5 | 0.06% | 0.05% | 5.7 | 3.40-8.63 | 989.3 | 460-1,783 |
| Altena | 11.2 | 0.07% | 0.12% | 10.5 | 6.78-15.78 | 736.5 | 250-1,258 |
| Ameland | 3.2 | 0.02% | 0.01% | 4.1 | 1.65-7.43 | 723.1 | 175-1,538 |
| Amersfoort | 155.6 | 0.93% | 0.97% | 6.3 | 5.22-8.01 | 979.7 | 389-1,889 |
| Amstelveen | 90.1 | 0.54% | 0.55% | 6.1 | 4.86-7.71 | 1,100.7 | 429-1,879 |
| Amsterdam | 860.2 | 5.16% | 5.91% | 7.0 | 6.12-8.05 | 1,139.3 | 433-2,269 |
| Apeldoorn | 161.4 | 0.97% | 0.89% | 5.6 | 4.52-6.84 | 1,027.9 | 356-2,174 |
| Appingedam | 11.2 | 0.07% | 0.05% | 4.1 | 2.07-8.05 | 551.7 | 166-1,026 |
| Arnhem | 158.3 | 0.95% | 0.94% | 6.0 | 4.68-7.35 | 1,205.9 | 436-2,035 |
| Assen | 67.4 | 0.40% | 0.33% | 4.9 | 3.47-7.16 | 723.0 | 295-1,464 |
| Asten | 16.3 | 0.10% | 0.10% | 6.3 | 3.91-10.86 | 930.3 | 439-1,746 |
| Baarle-Nassau | 6.4 | 0.04% | 0.03% | 5.3 | 2.72-9.07 | 992.8 | 356-1,911 |
| Baarn | 24.2 | 0.15% | 0.17% | 7.0 | 5.00-9.78 | 1,151.6 | 428-2,050 |
| Barendrecht | 48.0 | 0.29% | 0.31% | 6.6 | 5.00-8.62 | 1,252.9 | 543-2,065 |
| Barneveld | 57.3 | 0.34% | 0.35% | 6.1 | 4.74-7.80 | 1,023.4 | 405-1,744 |
| Beek | 15.3 | 0.09% | 0.08% | 5.5 | 3.60-9.04 | 927.8 | 348-1,613 |
| Beekdaelen | 11.2 | 0.07% | 0.05% | 4.8 | 2.78-7.85 | 866.3 | 0-1,589 |
| Beemster | 9.2 | 0.06% | 0.06% | 6.7 | 4.14-11.05 | 1,210.5 | 556-2,206 |
| Beesel | 13.0 | 0.08% | 0.07% | 5.6 | 3.12-9.65 | 830.9 | 287-1,651 |
| Berg en Dal | 34.3 | 0.21% | 0.21% | 6.3 | 4.49-9.30 | 1,047.0 | 362-2,143 |
| Bergeijk | 17.9 | 0.11% | 0.10% | 5.8 | 3.87-8.56 | 860.3 | 329-1,619 |
| Bergen (L.) | 12.6 | 0.08% | 0.07% | 5.7 | 3.40-8.96 | 806.6 | 302-1,675 |
| Bergen (NH.) | 29.4 | 0.18% | 0.16% | 5.4 | 3.80-7.85 | 928.5 | 330-1,623 |
| Bergen op Zoom | 66.1 | 0.40% | 0.36% | 5.6 | 3.98-9.82 | 764.2 | 222-1,438 |
| Berkelland | 43.4 | 0.26% | 0.23% | 5.3 | 3.70-7.38 | 860.2 | 270-1,849 |
| Bernheze | 30.3 | 0.18% | 0.20% | 6.6 | 4.59-8.50 | 1,077.1 | 497-1,706 |
| Best | 29.3 | 0.18% | 0.18% | 6.3 | 4.73-8.61 | 1,036.1 | 438-2,252 |
| Beuningen | 25.3 | 0.15% | 0.17% | 6.6 | 4.41-9.66 | 1,038.6 | 447-1,947 |
| Beverwijk | 40.6 | 0.24% | 0.25% | 6.3 | 4.92-8.26 | 1,148.5 | 468-2,392 |

Continued on next page

| Municipality | Population size (×1000) | Population percentage | Percentage of transmissions | Transmission risk | (90% range) | Seed Risk | (90% range) |
|---|---|---|---|---|---|---|---|
| Bladel | 19.8 | 0.12% | 0.11% | 5.7 | 3.86-7.91 | 1,048.5 | 484-1,738 |
| Blaricum | 10.9 | 0.07% | 0.07% | 6.7 | 4.02-11.18 | 1,084.1 | 473-1,790 |
| Bloemendaal | 22.8 | 0.14% | 0.15% | 6.8 | 5.00-9.62 | 1,278.9 | 603-2,072 |
| Bodegraven-Reeuwijk | 33.9 | 0.20% | 0.24% | 7.1 | 5.46-8.87 | 1,238.5 | 533-2,061 |
| Boekel | 10.2 | 0.06% | 0.07% | 6.5 | 3.90-10.63 | 1,269.9 | 538-2,464 |
| Borger-Odoorn | 24.9 | 0.15% | 0.10% | 4.2 | 2.60-6.90 | 527.2 | 155-1,014 |
| Borne | 22.7 | 0.14% | 0.11% | 4.9 | 2.90-6.91 | 773.5 | 243-1,626 |
| Borsele | 22.2 | 0.13% | 0.11% | 5.0 | 3.34-7.33 | 866.3 | 213-1,802 |
| Boxmeer | 28.4 | 0.17% | 0.17% | 6.0 | 4.03-10.25 | 1,070.9 | 457-1,872 |
| Boxtel | 30.1 | 0.18% | 0.19% | 6.3 | 4.59-8.42 | 1,054.8 | 522-2,078 |
| Breda | 183.4 | 1.10% | 1.11% | 6.2 | 4.98-7.88 | 1,237.6 | 508-2,123 |
| Brielle | 16.9 | 0.10% | 0.10% | 5.9 | 3.84-8.88 | 1,099.1 | 461-2,229 |
| Bronckhorst | 35.7 | 0.21% | 0.18% | 5.2 | 3.76-6.95 | 860.9 | 376-1,580 |
| Brummen | 20.3 | 0.12% | 0.12% | 5.8 | 3.88-9.06 | 971.7 | 435-1,543 |
| Brunssum | 27.6 | 0.17% | 0.15% | 5.5 | 3.62-8.14 | 780.1 | 357-1,660 |
| Bunnik | 14.6 | 0.09% | 0.10% | 6.6 | 4.73-10.13 | 1,135.8 | 520-2,069 |
| Bunschoten | 21.3 | 0.13% | 0.13% | 6.3 | 4.17-9.58 | 1,086.0 | 395-1,846 |
| Buren | 26.0 | 0.16% | 0.18% | 6.8 | 5.09-9.07 | 1,111.6 | 481-2,174 |
| Capelle aan den IJssel | 66.4 | 0.40% | 0.43% | 6.6 | 5.41-8.45 | 1,293.0 | 568-2,159 |
| Castricum | 35.3 | 0.21% | 0.22% | 6.4 | 4.77-8.40 | 1,142.8 | 385-2,017 |
| Coevorden | 34.6 | 0.21% | 0.15% | 4.3 | 2.68-6.70 | 634.6 | 167-1,203 |
| Cranendonck | 19.9 | 0.12% | 0.12% | 6.4 | 4.11-10.90 | 1,008.2 | 418-1,951 |
| Cuijk | 24.3 | 0.15% | 0.15% | 6.4 | 4.37-8.98 | 1,171.6 | 521-2,185 |
| Culemborg | 27.9 | 0.17% | 0.18% | 6.6 | 4.93-8.96 | 1,228.0 | 563-2,014 |
| Dalfsen | 27.9 | 0.17% | 0.15% | 5.5 | 3.59-9.82 | 757.6 | 228-1,535 |
| Dantumadiel | 18.5 | 0.11% | 0.09% | 4.9 | 3.02-7.96 | 804.8 | 327-1,478 |
| De Bilt | 42.2 | 0.25% | 0.26% | 6.2 | 4.81-8.52 | 1,175.8 | 423-2,181 |
| De Fryske Marren | 51.0 | 0.31% | 0.28% | 5.5 | 3.88-8.33 | 847.7 | 306-1,629 |
| De Ronde Venen | 43.6 | 0.26% | 0.28% | 6.6 | 4.89-8.79 | 1,241.8 | 575-2,166 |
| De Wolden | 23.4 | 0.14% | 0.12% | 5.0 | 3.29-7.46 | 915.8 | 405-1,848 |
| Delft | 102.7 | 0.62% | 0.72% | 7.1 | 5.98-8.51 | 1,142.7 | 565-2,134 |
| Delfzijl | 24.2 | 0.15% | 0.09% | 3.8 | 2.19-7.26 | 597.8 | 164-1,311 |
| Den Helder | 55.1 | 0.33% | 0.31% | 5.6 | 4.13-7.26 | 964.3 | 399-1,772 |
| Deurne | 31.8 | 0.19% | 0.19% | 6.0 | 4.27-8.40 | 921.9 | 282-1,740 |
| Deventer | 99.6 | 0.60% | 0.52% | 5.3 | 4.10-7.04 | 815.5 | 265-1,575 |
| Diemen | 28.6 | 0.17% | 0.23% | 8.1 | 6.11-11.25 | 1,088.1 | 487-2,068 |
| Dinkelland | 25.9 | 0.16% | 0.12% | 4.7 | 2.94-8.02 | 802.1 | 246-1,635 |
| Doesburg | 10.5 | 0.06% | 0.06% | 5.9 | 3.83-9.62 | 954.3 | 383-1,727 |
| Doetinchem | 57.2 | 0.34% | 0.31% | 5.5 | 3.95-7.16 | 846.0 | 349-1,751 |
| Dongen | 25.5 | 0.15% | 0.16% | 6.4 | 4.76-8.86 | 1,111.4 | 423-1,951 |
| Dordrecht | 118.1 | 0.71% | 0.73% | 6.3 | 5.11-7.59 | 1,182.2 | 479-2,126 |
| Drechterland | 19.0 | 0.11% | 0.12% | 6.2 | 3.79-8.95 | 1,162.7 | 404-2,099 |
| Drimmelen | 26.5 | 0.16% | 0.17% | 6.4 | 4.73-8.77 | 1,092.0 | 471-1,844 |
| Dronten | 40.2 | 0.24% | 0.24% | 6.1 | 4.39-8.42 | 968.3 | 313-1,785 |
| Druten | 18.3 | 0.11% | 0.12% | 6.5 | 4.41-9.48 | 1,154.3 | 513-2,052 |
| Duiven | 24.9 | 0.15% | 0.15% | 6.1 | 4.12-8.77 | 1,060.6 | 393-1,897 |
| Echt-Susteren | 31.1 | 0.19% | 0.17% | 5.6 | 3.99-8.96 | 932.8 | 308-1,706 |
| Edam-Volendam | 35.7 | 0.21% | 0.22% | 6.3 | 4.21-9.69 | 1,005.8 | 394-1,780 |
| Ede | 115.2 | 0.69% | 0.67% | 5.9 | 4.76-7.49 | 1,132.7 | 455-2,089 |
| Eemnes | 8.6 | 0.05% | 0.05% | 6.4 | 4.11-10.33 | 1,230.7 | 560-2,028 |
| Eersel | 18.6 | 0.11% | 0.12% | 6.5 | 4.41-8.83 | 1,141.4 | 532-2,211 |
| Eijsden-Margraten | 25.1 | 0.15% | 0.14% | 5.7 | 3.56-9.23 | 787.9 | 264-1,535 |
| Eindhoven | 230.5 | 1.38% | 1.43% | 6.3 | 5.10-7.66 | 994.6 | 375-1,879 |
| Elburg | 22.5 | 0.14% | 0.14% | 6.4 | 4.51-9.32 | 995.7 | 383-1,889 |
| Emmen | 106.4 | 0.64% | 0.46% | 4.4 | 3.25-6.09 | 626.3 | 118-1,452 |
| Enkhuizen | 18.0 | 0.11% | 0.11% | 6.4 | 4.29-8.98 | 1,151.0 | 516-1,980 |
| Enschede | 158.2 | 0.95% | 0.69% | 4.4 | 3.50-5.77 | 673.3 | 197-1,616 |
| Epe | 32.8 | 0.20% | 0.19% | 5.9 | 4.11-8.32 | 837.7 | 349-1,701 |
| Ermelo | 26.3 | 0.16% | 0.17% | 6.5 | 4.74-8.78 | 1,148.0 | 560-1,968 |
| Etten-Leur | 43.2 | 0.26% | 0.27% | 6.4 | 4.68-8.84 | 1,040.8 | 392-1,854 |
| Geertruidenberg | 21.0 | 0.13% | 0.14% | 6.6 | 4.88-8.75 | 1,177.9 | 494-2,162 |
| Geldrop-Mierlo | 39.1 | 0.23% | 0.24% | 6.3 | 4.63-8.45 | 1,216.7 | 438-2,256 |
| Gemert-Bakel | 30.0 | 0.18% | 0.19% | 6.3 | 4.53-9.74 | 838.4 | 271-1,539 |

Continued on next page

| Municipality | Population size (×1000) | Population percentage | Percentage of transmissions | Transmission risk | (90% range) | Seed Risk | (90% range) |
|---|---|---|---|---|---|---|---|
| Gennep | 16.7 | 0.10% | 0.09% | 5.6 | 3.82-9.11 | 1,046.3 | 507-2,015 |
| Gilze en Rijen | 25.9 | 0.16% | 0.16% | 6.3 | 4.49-9.53 | 973.4 | 371-2,055 |
| Goeree-Overflakkee | 49.2 | 0.30% | 0.27% | 5.5 | 3.89-7.86 | 960.2 | 388-1,780 |
| Goes | 37.1 | 0.22% | 0.18% | 4.9 | 3.26-7.46 | 754.9 | 261-1,601 |
| Goirle | 23.1 | 0.14% | 0.14% | 6.3 | 4.82-8.75 | 1,198.0 | 550-1,966 |
| Gooise Meren | 57.1 | 0.34% | 0.37% | 6.6 | 5.16-8.98 | 1,242.9 | 587-2,131 |
| Gorinchem | 35.9 | 0.22% | 0.25% | 7.0 | 5.18-9.96 | 1,194.6 | 507-2,088 |
| Gouda | 72.6 | 0.44% | 0.46% | 6.4 | 5.23-8.85 | 1,069.3 | 448-1,782 |
| Grave | 12.1 | 0.07% | 0.08% | 6.7 | 4.20-10.74 | 1,053.7 | 534-1,872 |
| Groningen | 203.2 | 1.22% | 1.00% | 5.0 | 3.74-6.80 | 733.7 | 192-1,389 |
| Gulpen-Wittem | 13.8 | 0.08% | 0.07% | 5.1 | 2.87-8.03 | 856.9 | 417-1,372 |
| Haaksbergen | 23.6 | 0.14% | 0.11% | 4.8 | 3.23-7.27 | 815.0 | 290-1,471 |
| Haaren | 13.6 | 0.08% | 0.09% | 6.5 | 4.57-10.26 | 1,038.5 | 435-1,706 |
| Haarlem | 160.5 | 0.96% | 1.02% | 6.4 | 5.53-7.51 | 1,202.1 | 564-2,033 |
| Haarlemmermeer | 147.3 | 0.88% | 0.97% | 6.7 | 5.30-8.28 | 1,239.2 | 691-2,148 |
| Halderberge | 29.8 | 0.18% | 0.17% | 5.6 | 3.79-8.00 | 1,001.7 | 349-2,267 |
| Hardenberg | 60.3 | 0.36% | 0.31% | 5.3 | 3.82-7.79 | 855.8 | 231-1,870 |
| Harderwijk | 46.7 | 0.28% | 0.28% | 6.0 | 4.44-8.52 | 981.6 | 403-1,720 |
| Hardinxveld-Giessendam | 17.5 | 0.11% | 0.11% | 6.5 | 4.30-9.65 | 1,183.0 | 463-2,271 |
| Harlingen | 15.3 | 0.09% | 0.08% | 5.0 | 2.99-8.41 | 825.9 | 347-1,621 |
| Hattem | 11.6 | 0.07% | 0.07% | 5.9 | 3.73-9.03 | 958.7 | 400-1,756 |
| Heemskerk | 38.8 | 0.23% | 0.28% | 7.3 | 5.42-9.41 | 1,312.5 | 535-2,247 |
| Heemstede | 26.8 | 0.16% | 0.18% | 6.9 | 5.39-10.07 | 1,283.3 | 564-2,394 |
| Heerde | 18.1 | 0.11% | 0.10% | 5.6 | 4.04-8.06 | 931.8 | 471-1,568 |
| Heerenveen | 49.6 | 0.30% | 0.26% | 5.3 | 3.75-7.44 | 811.7 | 272-1,624 |
| Heerhugowaard | 56.0 | 0.34% | 0.33% | 5.9 | 4.15-7.94 | 1,007.2 | 352-2,023 |
| Heerlen | 86.6 | 0.52% | 0.42% | 5.0 | 3.97-6.31 | 857.5 | 239-1,669 |
| Heeze-Leende | 15.5 | 0.09% | 0.09% | 6.1 | 3.85-9.04 | 988.9 | 449-1,646 |
| Heiloo | 23.0 | 0.14% | 0.14% | 6.2 | 4.58-9.30 | 1,103.3 | 444-1,861 |
| Hellendoorn | 35.4 | 0.21% | 0.19% | 5.4 | 3.72-8.57 | 774.4 | 363-1,353 |
| Hellevoetsluis | 39.6 | 0.24% | 0.25% | 6.3 | 4.17-10.43 | 989.3 | 401-1,713 |
| Helmond | 90.9 | 0.55% | 0.52% | 5.7 | 4.15-7.46 | 911.9 | 308-1,962 |
| Hendrik-Ido-Ambacht | 30.4 | 0.18% | 0.19% | 6.3 | 4.84-8.39 | 1,127.5 | 493-2,240 |
| Hengelo | 80.3 | 0.48% | 0.41% | 5.1 | 3.76-7.25 | 661.7 | 169-1,458 |
| Het Hogeland | 11.2 | 0.07% | 0.05% | 5.0 | 2.37-11.12 | 667.4 | 0-1,641 |
| Heumen | 16.0 | 0.10% | 0.09% | 5.8 | 3.88-8.59 | 1,031.5 | 385-1,772 |
| Heusden | 43.6 | 0.26% | 0.29% | 6.7 | 4.73-9.55 | 1,187.6 | 581-2,182 |
| Hillegom | 21.4 | 0.13% | 0.14% | 6.6 | 5.01-8.91 | 1,273.6 | 631-2,178 |
| Hilvarenbeek | 14.9 | 0.09% | 0.13% | 8.8 | 6.18-12.97 | 1,051.9 | 380-1,947 |
| Hilversum | 89.7 | 0.54% | 0.60% | 6.8 | 5.43-8.84 | 1,193.5 | 536-2,409 |
| Hoeksche Waard | 11.2 | 0.07% | 0.07% | 6.0 | 3.67-8.34 | 1,099.8 | 488-2,213 |
| Hof van Twente | 34.5 | 0.21% | 0.17% | 5.1 | 3.71-7.07 | 746.8 | 296-1,430 |
| Hollands Kroon | 46.9 | 0.28% | 0.26% | 5.5 | 3.81-8.42 | 821.0 | 261-1,549 |
| Hoogeveen | 55.0 | 0.33% | 0.28% | 5.1 | 3.45-7.81 | 732.8 | 232-1,468 |
| Hoorn | 72.6 | 0.44% | 0.44% | 6.1 | 4.82-8.21 | 1,016.8 | 430-1,746 |
| Horst aan de Maas | 41.7 | 0.25% | 0.23% | 5.7 | 4.06-8.10 | 939.7 | 392-1,756 |
| Houten | 49.5 | 0.30% | 0.32% | 6.6 | 4.85-8.52 | 1,160.5 | 419-1,911 |
| Huizen | 40.9 | 0.25% | 0.26% | 6.6 | 4.94-9.21 | 1,163.4 | 580-2,208 |
| Hulst | 27.0 | 0.16% | 0.14% | 5.4 | 3.62-9.05 | 849.4 | 245-1,519 |
| IJsselstein | 33.6 | 0.20% | 0.24% | 7.2 | 5.53-9.73 | 1,110.0 | 492-1,863 |
| Kaag en Braassem | 26.4 | 0.16% | 0.18% | 6.9 | 5.07-9.75 | 1,085.7 | 418-1,962 |
| Kampen | 53.2 | 0.32% | 0.29% | 5.5 | 4.28-6.96 | 968.9 | 310-1,856 |
| Kapelle | 12.4 | 0.07% | 0.06% | 4.7 | 2.96-7.22 | 760.3 | 284-1,560 |
| Katwijk | 64.6 | 0.39% | 0.45% | 7.1 | 5.16-9.42 | 1,213.1 | 590-1,936 |
| Kerkrade | 45.1 | 0.27% | 0.22% | 4.9 | 3.54-6.90 | 865.3 | 237-1,735 |
| Koggenland | 22.4 | 0.13% | 0.13% | 5.8 | 4.19-7.87 | 1,039.7 | 438-2,054 |
| Krimpen aan den IJssel | 28.8 | 0.17% | 0.21% | 7.4 | 5.85-9.18 | 1,505.7 | 743-2,535 |
| Krimpenerwaard | 55.5 | 0.33% | 0.34% | 6.2 | 4.68-7.93 | 1,174.8 | 556-1,971 |
| Laarbeek | 21.8 | 0.13% | 0.15% | 7.1 | 4.83-10.83 | 1,071.9 | 441-1,972 |
| Landerd | 14.8 | 0.09% | 0.09% | 6.3 | 3.95-10.00 | 1,108.1 | 428-1,827 |
| Landgraaf | 37.0 | 0.22% | 0.18% | 4.9 | 3.30-7.46 | 812.1 | 262-1,501 |
| Landsmeer | 11.0 | 0.07% | 0.08% | 7.1 | 4.62-10.13 | 1,251.3 | 479-2,146 |

Continued on next page

| Municipality | Population size (×1000) | Population percentage | Percentage of transmissions | Transmission risk | (90% range) | Seed Risk | (90% range) |
|---|---|---|---|---|---|---|---|
| Langedijk | 27.5 | 0.17% | 0.16% | 5.7 | 3.91-8.80 | 958.4 | 392-1,689 |
| Lansingerland | 61.1 | 0.37% | 0.40% | 6.7 | 5.13-8.82 | 1,266.2 | 594-2,148 |
| Laren | 10.7 | 0.06% | 0.07% | 6.6 | 4.49-9.13 | 1,245.4 | 562-2,276 |
| Leeuwarden | 122.4 | 0.73% | 0.61% | 5.1 | 3.85-6.88 | 772.9 | 316-1,613 |
| Leiden | 124.1 | 0.75% | 0.84% | 6.9 | 5.55-8.41 | 1,235.7 | 507-2,149 |
| Leiderdorp | 26.7 | 0.16% | 0.18% | 6.7 | 4.91-9.44 | 1,208.8 | 537-2,217 |
| Leidschendam-Voorburg | 74.8 | 0.45% | 0.49% | 6.6 | 5.47-8.38 | 1,285.7 | 670-2,274 |
| Lelystad | 77.6 | 0.47% | 0.44% | 5.8 | 4.38-7.85 | 971.8 | 347-1,934 |
| Leudal | 35.3 | 0.21% | 0.18% | 5.3 | 3.71-7.48 | 848.5 | 249-1,561 |
| Leusden | 29.2 | 0.18% | 0.19% | 6.7 | 4.62-10.72 | 1,337.6 | 669-2,601 |
| Lingewaard | 45.9 | 0.28% | 0.28% | 6.2 | 4.64-8.34 | 1,001.9 | 411-1,776 |
| Lisse | 22.4 | 0.13% | 0.16% | 7.0 | 5.06-9.17 | 1,213.7 | 542-2,307 |
| Lochem | 33.1 | 0.20% | 0.18% | 5.5 | 3.61-8.04 | 808.0 | 349-1,550 |
| Loon op Zand | 22.6 | 0.14% | 0.14% | 6.4 | 4.94-8.16 | 1,229.2 | 574-2,166 |
| Lopik | 13.9 | 0.08% | 0.10% | 7.0 | 4.99-10.35 | 1,050.6 | 418-2,025 |
| Loppersum | 9.0 | 0.05% | 0.04% | 4.0 | 2.12-7.09 | 744.7 | 220-1,631 |
| Losser | 22.2 | 0.13% | 0.09% | 4.2 | 2.42-8.42 | 640.5 | 117-1,599 |
| Maasdriel | 24.0 | 0.14% | 0.17% | 7.1 | 4.65-10.36 | 1,027.9 | 466-1,770 |
| Maasgouw | 23.2 | 0.14% | 0.13% | 5.7 | 3.75-7.91 | 776.3 | 300-1,326 |
| Maassluis | 32.2 | 0.19% | 0.22% | 6.8 | 4.78-9.04 | 1,094.9 | 428-1,925 |
| Maastricht | 121.7 | 0.73% | 0.60% | 5.0 | 3.89-6.58 | 745.7 | 276-1,410 |
| Medemblik | 44.2 | 0.27% | 0.24% | 5.5 | 4.07-7.51 | 993.9 | 361-1,899 |
| Meerssen | 18.5 | 0.11% | 0.10% | 5.5 | 3.34-8.20 | 928.5 | 302-1,767 |
| Meierijstad | 80.3 | 0.48% | 0.50% | 6.3 | 5.11-8.08 | 1,130.5 | 543-1,946 |
| Meppel | 33.0 | 0.20% | 0.17% | 5.3 | 3.96-7.56 | 858.9 | 338-1,636 |
| Middelburg | 48.3 | 0.29% | 0.20% | 4.3 | 2.93-6.11 | 691.2 | 171-1,499 |
| Midden-Delfland | 19.1 | 0.11% | 0.16% | 8.7 | 6.34-12.85 | 1,178.7 | 483-1,868 |
| Midden-Drenthe | 32.5 | 0.20% | 0.17% | 5.2 | 3.37-9.10 | 678.1 | 183-1,351 |
| Midden-Groningen | 60.5 | 0.36% | 0.26% | 4.4 | 3.23-6.84 | 726.7 | 119-1,523 |
| Mill en Sint Hubert | 10.5 | 0.06% | 0.07% | 6.6 | 4.37-9.89 | 1,035.5 | 438-1,777 |
| Moerdijk | 36.6 | 0.22% | 0.22% | 6.2 | 4.76-8.07 | 1,102.0 | 474-1,957 |
| Molenlanden | 11.2 | 0.07% | 0.07% | 6.8 | 4.43-10.74 | 1,289.2 | 444-2,627 |
| Montferland | 35.3 | 0.21% | 0.19% | 5.6 | 4.13-7.46 | 840.1 | 254-1,492 |
| Montfoort | 13.4 | 0.08% | 0.09% | 7.0 | 4.30-10.58 | 1,170.0 | 535-2,120 |
| Mook en Middelaar | 7.3 | 0.04% | 0.04% | 5.7 | 3.39-8.19 | 1,004.1 | 448-1,802 |
| Neder-Betuwe | 23.3 | 0.14% | 0.14% | 6.1 | 4.24-8.88 | 1,149.7 | 368-2,188 |
| Nederweert | 16.6 | 0.10% | 0.10% | 6.1 | 4.06-9.03 | 974.9 | 338-1,746 |
| Nieuwegein | 62.5 | 0.38% | 0.42% | 6.9 | 5.63-8.60 | 1,327.6 | 656-2,282 |
| Nieuwkoop | 28.0 | 0.17% | 0.19% | 6.7 | 4.98-8.89 | 1,152.1 | 449-2,115 |
| Nijkerk | 42.3 | 0.25% | 0.26% | 6.3 | 4.69-8.23 | 1,052.0 | 496-1,831 |
| Nijmegen | 176.6 | 1.06% | 1.06% | 6.1 | 4.90-7.86 | 993.8 | 378-1,757 |
| Nissewaard | 84.2 | 0.51% | 0.50% | 6.0 | 4.63-7.92 | 1,011.1 | 473-1,790 |
| Noardeast-FryslÃ¢n | 11.2 | 0.07% | 0.04% | 4.0 | 2.26-7.00 | 663.1 | 150-1,300 |
| Noord-Beveland | 6.7 | 0.04% | 0.03% | 4.3 | 2.18-6.88 | 724.5 | 162-1,626 |
| Noordenveld | 30.8 | 0.18% | 0.14% | 4.7 | 3.16-7.10 | 717.9 | 247-1,432 |
| Noordoostpolder | 46.3 | 0.28% | 0.25% | 5.5 | 4.08-7.92 | 781.0 | 244-1,505 |
| Noordwijk | 25.8 | 0.15% | 0.17% | 6.6 | 4.46-10.03 | 1,163.7 | 507-2,023 |
| Nuenen, Gerwen en Nederwetten | 22.7 | 0.14% | 0.13% | 6.0 | 4.49-9.17 | 1,040.3 | 353-1,999 |
| Nunspeet | 27.1 | 0.16% | 0.16% | 6.2 | 4.27-10.36 | 1,051.6 | 418-1,923 |
| Oegstgeest | 23.9 | 0.14% | 0.19% | 8.2 | 5.81-11.07 | 1,228.3 | 560-2,151 |
| Oirschot | 18.1 | 0.11% | 0.11% | 6.4 | 4.16-9.40 | 1,178.4 | 482-2,202 |
| Oisterwijk | 25.4 | 0.15% | 0.16% | 6.3 | 3.89-10.61 | 1,023.6 | 303-1,820 |
| Oldambt | 37.5 | 0.23% | 0.16% | 4.3 | 2.55-7.58 | 488.4 | 111-1,109 |
| Oldebroek | 23.1 | 0.14% | 0.14% | 6.1 | 4.37-8.89 | 942.6 | 351-1,877 |
| Oldenzaal | 31.1 | 0.19% | 0.14% | 4.5 | 2.91-6.32 | 741.6 | 241-1,449 |
| Olst-Wijhe | 17.6 | 0.11% | 0.10% | 5.8 | 3.78-8.24 | 1,011.2 | 349-1,844 |
| Ommen | 17.4 | 0.10% | 0.10% | 5.7 | 3.14-10.47 | 885.3 | 259-1,651 |
| Oost Gelre | 29.2 | 0.18% | 0.14% | 5.0 | 3.36-7.15 | 747.0 | 202-1,678 |
| Oosterhout | 54.7 | 0.33% | 0.35% | 6.4 | 5.19-8.93 | 1,199.1 | 459-2,204 |
| Ooststellingwerf | 24.9 | 0.15% | 0.13% | 5.2 | 3.11-8.15 | 765.5 | 300-1,392 |

Continued on next page

| Municipality | Population size (×1000) | Population percentage | Percentage of transmissions | Transmission risk | (90% range) | Seed Risk | (90% range) |
|---|---|---|---|---|---|---|---|
| Oostzaan | 9.2 | 0.06% | 0.06% | 7.2 | 3.97-10.35 | 1,238.8 | 625-2,114 |
| Opmeer | 11.2 | 0.07% | 0.07% | 6.4 | 3.73-13.48 | 1,172.8 | 497-1,972 |
| Opsterland | 29.4 | 0.18% | 0.15% | 5.1 | 3.22-7.79 | 807.8 | 244-1,595 |
| Oss | 91.0 | 0.55% | 0.54% | 6.0 | 4.70-7.70 | 1,123.6 | 454-2,278 |
| Oude IJsselstreek | 38.8 | 0.23% | 0.20% | 5.1 | 3.39-7.50 | 815.5 | 198-1,549 |
| Ouder-Amstel | 13.3 | 0.08% | 0.10% | 7.8 | 5.37-10.70 | 1,338.0 | 538-2,237 |
| Oudewater | 9.7 | 0.06% | 0.07% | 7.3 | 4.58-11.03 | 1,281.6 | 659-2,141 |
| Overbetuwe | 47.0 | 0.28% | 0.28% | 5.9 | 4.62-7.43 | 1,107.5 | 427-2,135 |
| Papendrecht | 31.8 | 0.19% | 0.20% | 6.2 | 4.78-8.27 | 1,195.1 | 512-2,212 |
| Peel en Maas | 42.9 | 0.26% | 0.24% | 5.7 | 3.93-8.81 | 885.0 | 201-1,590 |
| Pekela | 11.9 | 0.07% | 0.05% | 4.4 | 2.18-7.13 | 613.1 | 197-1,246 |
| Pijnacker-Nootdorp | 53.6 | 0.32% | 0.42% | 8.0 | 6.25-10.82 | 1,333.9 | 495-2,415 |
| Purmerend | 79.6 | 0.48% | 0.50% | 6.4 | 4.75-8.85 | 1,105.3 | 403-2,124 |
| Putten | 23.8 | 0.14% | 0.16% | 6.8 | 4.93-9.18 | 1,192.0 | 491-2,243 |
| Raalte | 36.9 | 0.22% | 0.20% | 5.4 | 3.74-7.77 | 917.1 | 305-1,819 |
| Reimerswaal | 22.2 | 0.13% | 0.11% | 5.1 | 3.18-8.06 | 825.2 | 277-1,754 |
| Renkum | 30.9 | 0.19% | 0.20% | 6.5 | 4.60-9.59 | 1,154.6 | 399-2,208 |
| Renswoude | 4.6 | 0.03% | 0.04% | 8.2 | 3.48-20.76 | 1,196.4 | 538-1,915 |
| Reusel-De Mierden | 12.6 | 0.08% | 0.07% | 6.0 | 3.44-10.25 | 1,013.1 | 436-1,842 |
| Rheden | 43.3 | 0.26% | 0.24% | 5.7 | 4.36-8.35 | 839.5 | 264-1,543 |
| Rhenen | 19.4 | 0.12% | 0.14% | 7.3 | 5.25-10.05 | 1,199.9 | 576-2,257 |
| Ridderkerk | 45.7 | 0.27% | 0.28% | 6.2 | 4.78-8.72 | 1,069.9 | 487-2,018 |
| Rijssen-Holten | 37.8 | 0.23% | 0.20% | 5.4 | 3.71-7.88 | 788.5 | 199-1,539 |
| Rijswijk | 52.7 | 0.32% | 0.37% | 7.1 | 5.70-9.18 | 1,168.9 | 636-1,835 |
| Roerdalen | 20.1 | 0.12% | 0.10% | 5.2 | 3.38-7.84 | 953.6 | 356-1,776 |
| Roermond | 57.4 | 0.34% | 0.30% | 5.4 | 3.77-7.35 | 934.1 | 388-1,828 |
| Roosendaal | 76.6 | 0.46% | 0.44% | 5.8 | 4.55-7.31 | 1,006.3 | 378-1,840 |
| Rotterdam | 641.3 | 3.85% | 4.43% | 7.0 | 6.27-7.84 | 1,150.6 | 542-1,914 |
| Rozendaal | 1.3 | 0.01% | 0.00% | 3.5 | 1.13-6.78 | 1,049.5 | 443-1,678 |
| Rucphen | 22.2 | 0.13% | 0.14% | 6.5 | 4.25-10.06 | 1,063.5 | 401-1,955 |
| Schagen | 45.9 | 0.28% | 0.28% | 6.3 | 4.42-10.77 | 1,222.3 | 484-2,118 |
| Scherpenzeel | 9.5 | 0.06% | 0.06% | 6.2 | 4.08-9.62 | 1,029.1 | 444-1,881 |
| Schiedam | 77.4 | 0.46% | 0.60% | 7.9 | 6.35-9.66 | 1,162.2 | 449-1,983 |
| Schiermonnikoog | 0.5 | 0.00% | 0.00% | 2.8 | 0.28-7.26 | 1,014.5 | 413-1,872 |
| Schouwen-Duiveland | 33.3 | 0.20% | 0.16% | 4.8 | 3.47-6.76 | 860.2 | 267-1,942 |
| Simpelveld | 9.9 | 0.06% | 0.05% | 5.1 | 2.80-8.59 | 809.3 | 266-1,589 |
| Sint Anthonis | 11.2 | 0.07% | 0.07% | 6.0 | 3.67-9.44 | 1,049.3 | 371-2,036 |
| Sint-Michielsgestel | 28.5 | 0.17% | 0.19% | 6.9 | 5.06-9.86 | 1,148.4 | 446-1,936 |
| Sittard-Geleen | 92.2 | 0.55% | 0.47% | 5.2 | 3.98-6.84 | 819.5 | 207-1,702 |
| Sliedrecht | 24.6 | 0.15% | 0.17% | 6.8 | 4.97-10.17 | 1,171.2 | 577-2,176 |
| Sluis | 23.0 | 0.14% | 0.09% | 3.9 | 2.30-5.84 | 736.3 | 151-1,528 |
| Smallingerland | 55.4 | 0.33% | 0.29% | 5.3 | 3.82-7.45 | 749.8 | 224-1,434 |
| Soest | 45.6 | 0.27% | 0.28% | 6.3 | 4.93-7.89 | 1,237.5 | 508-2,158 |
| Someren | 18.8 | 0.11% | 0.12% | 6.3 | 4.14-8.87 | 844.7 | 297-1,442 |
| Son en Breugel | 16.2 | 0.10% | 0.10% | 6.5 | 4.70-9.07 | 1,069.1 | 525-1,875 |
| Stadskanaal | 31.4 | 0.19% | 0.14% | 4.5 | 2.65-7.55 | 641.7 | 124-1,456 |
| Staphorst | 16.4 | 0.10% | 0.08% | 5.2 | 3.22-7.54 | 735.5 | 303-1,486 |
| Stede Broec | 21.3 | 0.13% | 0.13% | 6.0 | 4.16-8.55 | 911.3 | 329-1,735 |
| Steenbergen | 25.0 | 0.15% | 0.15% | 6.0 | 3.83-10.11 | 740.7 | 274-1,223 |
| Steenwijkerland | 43.3 | 0.26% | 0.24% | 5.7 | 4.18-8.49 | 904.9 | 412-1,730 |
| Stein | 24.4 | 0.15% | 0.13% | 5.6 | 3.76-9.74 | 902.3 | 339-1,522 |
| Stichtse Vecht | 63.9 | 0.38% | 0.40% | 6.3 | 4.81-8.67 | 1,096.4 | 424-1,841 |
| SÃºdwest-FryslÃ¢n | 89.0 | 0.53% | 0.45% | 5.1 | 3.89-7.02 | 847.9 | 220-1,801 |
| Terneuzen | 54.1 | 0.32% | 0.25% | 4.7 | 3.01-6.39 | 782.6 | 251-1,640 |
| Terschelling | 4.5 | 0.03% | 0.02% | 4.8 | 1.95-11.39 | 738.0 | 228-1,485 |
| Texel | 13.0 | 0.08% | 0.07% | 5.3 | 2.98-8.39 | 890.4 | 300-1,881 |
| Teylingen | 36.5 | 0.22% | 0.23% | 6.5 | 4.89-9.40 | 1,236.0 | 623-1,961 |
| Tholen | 25.2 | 0.15% | 0.15% | 6.0 | 3.88-9.57 | 916.9 | 271-1,933 |
| Tiel | 41.5 | 0.25% | 0.25% | 6.1 | 4.85-7.79 | 1,152.1 | 500-2,245 |
| Tilburg | 216.5 | 1.30% | 1.28% | 6.0 | 4.83-7.36 | 1,133.0 | 511-2,083 |
| Tubbergen | 20.6 | 0.12% | 0.11% | 5.2 | 3.34-8.08 | 773.2 | 290-1,603 |
| Twenterand | 33.6 | 0.20% | 0.17% | 5.0 | 3.43-7.88 | 789.9 | 239-1,420 |

Continued on next page

| Municipality | Population size (×1000) | Population percentage | Percentage of transmissions | Transmission risk | (90% range) | Seed Risk | (90% range) |
|---|---|---|---|---|---|---|---|
| Tynaarlo | 33.2 | 0.20% | 0.14% | 4.2 | 2.65-6.17 | 590.4 | 186-1,174 |
| Tytsjerksteradiel | 31.3 | 0.19% | 0.16% | 5.1 | 3.38-7.38 | 832.9 | 309-1,547 |
| Uden | 41.2 | 0.25% | 0.25% | 6.2 | 4.60-8.53 | 962.3 | 292-1,657 |
| Uitgeest | 13.0 | 0.08% | 0.09% | 7.2 | 4.58-10.22 | 1,095.3 | 395-2,166 |
| Uithoorn | 28.9 | 0.17% | 0.19% | 6.7 | 4.85-9.58 | 1,185.1 | 485-2,122 |
| Urk | 20.2 | 0.12% | 0.13% | 6.4 | 3.95-9.79 | 906.3 | 282-1,848 |
| Utrecht | 350.4 | 2.10% | 2.33% | 6.7 | 5.75-7.91 | 1,080.2 | 520-1,941 |
| Utrechtse Heuvel-rug | 49.0 | 0.29% | 0.29% | 6.0 | 4.75-7.71 | 1,016.1 | 347-1,976 |
| Vaals | 9.6 | 0.06% | 0.05% | 5.2 | 3.05-8.09 | 881.8 | 324-1,752 |
| Valkenburg aan de Geul | 16.0 | 0.10% | 0.08% | 4.8 | 3.00-7.06 | 883.3 | 312-1,712 |
| Valkenswaard | 30.4 | 0.18% | 0.18% | 6.1 | 4.51-8.61 | 1,044.5 | 399-1,792 |
| Veendam | 26.9 | 0.16% | 0.12% | 4.5 | 2.60-6.93 | 692.9 | 150-1,509 |
| Veenendaal | 64.9 | 0.39% | 0.39% | 6.1 | 4.89-7.68 | 1,146.8 | 525-2,351 |
| Veere | 21.2 | 0.13% | 0.10% | 4.6 | 2.65-8.62 | 626.1 | 177-1,334 |
| Veldhoven | 44.6 | 0.27% | 0.26% | 5.8 | 4.42-7.40 | 906.7 | 434-1,709 |
| Velsen | 67.7 | 0.41% | 0.42% | 6.2 | 5.02-7.76 | 1,141.4 | 427-2,014 |
| Venlo | 101.0 | 0.61% | 0.53% | 5.3 | 4.06-7.40 | 781.1 | 256-1,449 |
| Venray | 42.8 | 0.26% | 0.23% | 5.4 | 3.93-7.34 | 852.3 | 178-1,644 |
| Vijfheerenlanden | 11.2 | 0.07% | 0.07% | 6.8 | 4.50-9.71 | 1,325.7 | 546-2,449 |
| Vlaardingen | 71.8 | 0.43% | 0.52% | 7.4 | 5.69-9.44 | 1,001.6 | 349-1,746 |
| Vlieland | 0.6 | 0.00% | 0.00% | 2.5 | 0.29-5.93 | 991.8 | 352-1,828 |
| Vlissingen | 44.1 | 0.26% | 0.19% | 4.4 | 2.92-7.06 | 570.9 | 161-1,355 |
| Voerendaal | 12.0 | 0.07% | 0.06% | 5.2 | 3.17-7.51 | 919.1 | 272-1,708 |
| Voorschoten | 25.0 | 0.15% | 0.17% | 6.8 | 5.05-10.65 | 1,146.1 | 576-1,929 |
| Voorst | 23.9 | 0.14% | 0.13% | 5.7 | 3.93-7.53 | 899.9 | 338-1,672 |
| Vught | 26.0 | 0.16% | 0.17% | 6.5 | 4.93-9.59 | 1,184.7 | 579-1,994 |
| Waadhoeke | 45.6 | 0.27% | 0.24% | 5.3 | 3.79-7.69 | 806.4 | 265-1,556 |
| Waalre | 16.9 | 0.10% | 0.11% | 6.6 | 4.23-9.66 | 1,163.8 | 554-2,136 |
| Waalwijk | 47.7 | 0.29% | 0.28% | 6.0 | 4.59-8.05 | 1,035.3 | 345-1,939 |
| Waddinxveen | 27.9 | 0.17% | 0.19% | 7.1 | 5.01-9.23 | 1,197.4 | 579-2,170 |
| Wageningen | 38.1 | 0.23% | 0.25% | 6.6 | 5.22-8.14 | 1,066.0 | 414-1,870 |
| Wassenaar | 25.6 | 0.15% | 0.22% | 8.5 | 6.60-11.25 | 1,047.9 | 504-1,734 |
| Waterland | 16.8 | 0.10% | 0.12% | 7.0 | 5.10-9.29 | 1,160.7 | 370-1,920 |
| Weert | 49.1 | 0.29% | 0.25% | 5.2 | 4.00-7.02 | 771.5 | 283-1,371 |
| Weesp | 18.8 | 0.11% | 0.13% | 7.2 | 5.13-10.23 | 1,127.9 | 567-1,909 |
| West Betuwe | 11.2 | 0.07% | 0.08% | 6.8 | 4.45-9.77 | 1,173.8 | 397-2,128 |
| West Maas en Waal | 18.5 | 0.11% | 0.12% | 6.5 | 4.48-10.67 | 1,036.1 | 411-1,738 |
| Westerkwartier | 11.2 | 0.07% | 0.05% | 4.6 | 2.18-9.95 | 542.8 | 184-927 |
| Westerveld | 18.9 | 0.11% | 0.08% | 4.5 | 3.00-7.08 | 734.0 | 241-1,331 |
| Westervoort | 14.5 | 0.09% | 0.09% | 6.0 | 4.19-8.41 | 1,001.4 | 482-1,824 |
| Westerwolde | 24.8 | 0.15% | 0.10% | 4.0 | 2.52-7.51 | 555.6 | 102-1,278 |
| Westland | 108.3 | 0.65% | 0.64% | 6.0 | 4.95-7.47 | 1,151.6 | 502-2,217 |
| Weststellingwerf | 25.4 | 0.15% | 0.14% | 5.4 | 3.55-7.99 | 856.4 | 250-1,631 |
| Westvoorne | 14.3 | 0.09% | 0.08% | 5.6 | 3.43-8.50 | 830.4 | 268-1,603 |
| Wierden | 23.9 | 0.14% | 0.14% | 5.8 | 3.34-8.98 | 813.0 | 312-1,476 |
| Wijchen | 40.4 | 0.24% | 0.25% | 6.3 | 4.79-8.63 | 1,170.1 | 543-2,077 |
| Wijdemeren | 23.4 | 0.14% | 0.16% | 7.1 | 5.24-10.73 | 1,229.1 | 530-2,011 |
| Wijk bij Duurst-ede | 23.3 | 0.14% | 0.16% | 6.8 | 4.61-9.56 | 1,152.4 | 497-2,046 |
| Winterswijk | 28.5 | 0.17% | 0.14% | 5.2 | 3.38-8.43 | 862.3 | 244-1,715 |
| Woensdrecht | 21.4 | 0.13% | 0.12% | 5.8 | 3.92-9.45 | 1,020.6 | 304-1,715 |
| Woerden | 51.5 | 0.31% | 0.33% | 6.4 | 4.94-8.65 | 1,061.6 | 459-1,848 |
| Wormerland | 15.8 | 0.09% | 0.12% | 7.5 | 5.13-12.24 | 1,322.7 | 722-2,158 |
| Woudenberg | 12.6 | 0.08% | 0.09% | 7.4 | 4.33-15.09 | 1,126.5 | 478-2,008 |
| Zaanstad | 155.1 | 0.93% | 1.00% | 6.6 | 5.45-8.20 | 1,166.7 | 497-1,957 |
| Zaltbommel | 27.9 | 0.17% | 0.18% | 6.5 | 4.86-9.27 | 1,170.1 | 427-2,151 |
| Zandvoort | 16.3 | 0.10% | 0.11% | 6.7 | 4.38-9.29 | 1,075.2 | 480-1,796 |
| Zeewolde | 21.8 | 0.13% | 0.15% | 6.8 | 4.67-10.15 | 1,096.6 | 479-1,947 |
| Zeist | 63.6 | 0.38% | 0.39% | 6.2 | 4.75-7.69 | 1,082.6 | 502-2,059 |
| Zevenaar | 43.0 | 0.26% | 0.23% | 5.3 | 4.11-6.84 | 937.2 | 279-1,746 |
| Zoetermeer | 124.4 | 0.75% | 0.81% | 6.6 | 5.41-8.03 | 1,301.1 | 576-2,109 |
| Zoeterwoude | 8.1 | 0.05% | 0.06% | 7.1 | 4.62-10.28 | 1,235.4 | 541-2,168 |
| Zuidplas | 42.1 | 0.25% | 0.29% | 7.0 | 5.61-9.17 | 1,261.0 | 590-2,465 |

Continued on next page

| Municipality | Population size (×1000) | Population percentage | Percentage of transmissions | Transmission risk | (90% range) | Seed Risk | (90% range) |
|---|---|---|---|---|---|---|---|
| Zundert | 21.4 | 0.13% | 0.14% | 6.5 | 4.65-8.87 | 1,155.6 | 564-1,985 |
| Zutphen | 47.0 | 0.28% | 0.25% | 5.4 | 3.90-7.11 | 892.7 | 307-1,682 |
| Zwartewaterland | 22.0 | 0.13% | 0.13% | 5.9 | 3.65-8.05 | 906.8 | 294-1,699 |
| Zwijndrecht | 44.3 | 0.27% | 0.29% | 6.6 | 4.69-8.81 | 1,127.5 | 541-1,957 |
| Zwolle | 126.6 | 0.76% | 0.66% | 5.3 | 4.16-7.20 | 851.0 | 270-1,530 |

**Supplementary Table SI.3:** The full list of population (data from 2019) and 21-days transmission risk scores and seed risk scores of the municipalities in the Netherlands after an initial introduction in students.

| Municipality | Population size (×1000) | Population percentage | Percentage of transmissions | Transmission risk | (90% range) | Seed Risk | (90% range) |
|---|---|---|---|---|---|---|---|
| 's-Gravenhage | 535.6 | 3.22% | 3.71% | 22.8 | 20.76-25.01 | 3,511.1 | 1,695-6,840 |
| 's-Hertogenbosch | 153.5 | 0.92% | 1.00% | 21.4 | 18.52-24.76 | 3,456.9 | 1,613-5,806 |
| Aa en Hunze | 24.8 | 0.15% | 0.12% | 15.3 | 10.52-23.92 | 2,050.1 | 630-3,771 |
| Aalsmeer | 31.0 | 0.19% | 0.20% | 20.9 | 16.87-25.79 | 4,160.0 | 1,893-8,547 |
| Aalten | 26.5 | 0.16% | 0.12% | 15.2 | 10.78-20.89 | 2,069.8 | 551-4,879 |
| Achtkarspelen | 27.5 | 0.17% | 0.13% | 16.0 | 12.23-24.12 | 2,053.1 | 607-4,102 |
| Alblasserdam | 19.7 | 0.12% | 0.13% | 21.0 | 16.38-25.36 | 3,975.7 | 1,882-7,149 |
| Albrandswaard | 24.8 | 0.15% | 0.15% | 20.5 | 16.67-27.65 | 3,682.8 | 1,702-6,507 |
| Alkmaar | 108.2 | 0.65% | 0.64% | 19.4 | 16.82-22.84 | 3,507.1 | 1,317-6,567 |
| Almelo | 72.4 | 0.43% | 0.35% | 16.0 | 12.65-20.06 | 2,536.6 | 785-5,629 |
| Almere | 206.2 | 1.24% | 1.40% | 22.3 | 19.85-25.52 | 3,704.0 | 1,702-6,266 |
| Alphen aan den Rijn | 110.2 | 0.66% | 0.76% | 22.7 | 19.82-26.49 | 3,649.6 | 2,176-5,930 |
| Alphen-Chaam | 9.5 | 0.06% | 0.05% | 18.5 | 13.15-27.52 | 3,216.2 | 1,479-5,905 |
| Altena | 11.2 | 0.07% | 0.10% | 30.1 | 23.43-41.75 | 2,403.1 | 855-4,086 |
| Ameland | 3.2 | 0.02% | 0.01% | 13.0 | 7.36-20.64 | 2,343.2 | 612-4,972 |
| Amersfoort | 155.6 | 0.93% | 1.02% | 21.7 | 19.04-25.96 | 3,179.7 | 1,248-6,253 |
| Amstelveen | 90.1 | 0.54% | 0.57% | 20.8 | 17.92-24.14 | 3,568.5 | 1,322-6,085 |
| Amsterdam | 860.2 | 5.16% | 6.20% | 23.8 | 21.31-26.59 | 3,717.7 | 1,431-7,581 |
| Apeldoorn | 161.4 | 0.97% | 0.95% | 19.3 | 16.43-22.12 | 3,317.7 | 1,177-7,114 |
| Appingedam | 11.2 | 0.07% | 0.05% | 13.4 | 8.61-21.50 | 1,790.9 | 580-3,494 |
| Arnhem | 158.3 | 0.95% | 0.98% | 20.5 | 17.31-24.16 | 3,900.3 | 1,398-6,744 |
| Assen | 67.4 | 0.40% | 0.33% | 16.2 | 12.18-21.10 | 2,345.6 | 1,045-4,593 |
| Asten | 16.3 | 0.10% | 0.09% | 18.8 | 14.23-27.32 | 3,012.7 | 1,412-5,552 |
| Baarle-Nassau | 6.4 | 0.04% | 0.03% | 16.3 | 10.94-25.45 | 3,212.7 | 1,090-6,224 |
| Baarn | 24.2 | 0.15% | 0.15% | 20.5 | 15.74-26.82 | 3,752.3 | 1,397-6,699 |
| Barendrecht | 48.0 | 0.29% | 0.30% | 20.9 | 17.42-25.73 | 4,078.6 | 1,704-6,786 |
| Barneveld | 57.3 | 0.34% | 0.33% | 19.2 | 15.69-23.34 | 3,344.9 | 1,299-5,729 |
| Beek | 15.3 | 0.09% | 0.08% | 17.1 | 13.16-23.60 | 2,995.8 | 1,146-5,285 |
| Beekdaelen | 11.2 | 0.07% | 0.05% | 15.9 | 11.05-23.28 | 2,817.3 | 0-5,379 |
| Beemster | 9.2 | 0.06% | 0.06% | 20.4 | 14.98-27.85 | 3,932.0 | 1,816-7,062 |
| Beesel | 13.0 | 0.08% | 0.07% | 16.9 | 12.03-23.82 | 2,669.2 | 917-5,094 |
| Berg en Dal | 34.3 | 0.21% | 0.20% | 19.2 | 14.91-26.77 | 3,388.8 | 1,156-6,690 |
| Bergeijk | 17.9 | 0.11% | 0.10% | 17.6 | 13.13-22.01 | 2,796.1 | 1,102-5,132 |
| Bergen (L.) | 12.6 | 0.08% | 0.07% | 17.6 | 12.34-23.17 | 2,602.0 | 989-5,465 |
| Bergen (NH.) | 29.4 | 0.18% | 0.15% | 16.8 | 12.98-22.81 | 3,035.4 | 1,089-5,254 |
| Bergen op Zoom | 66.1 | 0.40% | 0.35% | 17.6 | 14.49-25.49 | 2,485.0 | 749-4,707 |
| Berkelland | 43.4 | 0.26% | 0.22% | 16.8 | 13.31-22.04 | 2,779.4 | 891-5,928 |
| Bernheze | 30.3 | 0.18% | 0.18% | 19.4 | 15.56-23.76 | 3,492.5 | 1,599-5,561 |
| Best | 29.3 | 0.18% | 0.17% | 19.0 | 15.19-24.12 | 3,334.7 | 1,364-7,036 |
| Beuningen | 25.3 | 0.15% | 0.15% | 20.1 | 15.34-26.18 | 3,357.3 | 1,445-6,508 |
| Beverwijk | 40.6 | 0.24% | 0.24% | 19.5 | 16.29-23.92 | 3,756.0 | 1,610-8,002 |
| Bladel | 19.8 | 0.12% | 0.11% | 17.7 | 13.80-23.47 | 3,381.9 | 1,629-5,698 |
| Blaricum | 10.9 | 0.07% | 0.07% | 20.8 | 14.77-30.14 | 3,511.1 | 1,486-5,921 |
| Bloemendaal | 22.8 | 0.14% | 0.14% | 20.4 | 16.58-27.29 | 4,152.1 | 1,962-6,625 |
| Bodegraven-Reeuwijk | 33.9 | 0.20% | 0.22% | 21.6 | 17.55-26.55 | 4,021.3 | 1,759-6,736 |
| Boekel | 10.2 | 0.06% | 0.06% | 19.8 | 14.65-27.00 | 4,113.8 | 1,772-7,966 |
| Borger-Odoorn | 24.9 | 0.15% | 0.10% | 13.8 | 9.99-19.57 | 1,713.4 | 540-3,286 |
| Borne | 22.7 | 0.14% | 0.11% | 15.3 | 10.91-21.16 | 2,482.8 | 830-5,209 |
| Borsele | 22.2 | 0.13% | 0.11% | 16.0 | 12.15-21.51 | 2,803.9 | 722-5,976 |
| Boxmeer | 28.4 | 0.17% | 0.16% | 18.5 | 13.79-27.04 | 3,454.1 | 1,513-6,148 |
| Boxtel | 30.1 | 0.18% | 0.17% | 18.9 | 15.16-24.11 | 3,421.9 | 1,721-6,685 |

Continued on next page

| Municipality | Population size (×1000) | Population percentage | Percentage of transmissions | Transmission risk | (90% range) | Seed Risk | (90% range) |
|---|---|---|---|---|---|---|---|
| Breda | 183.4 | 1.10% | 1.19% | 21.4 | 18.40-25.60 | 4,043.4 | 1,686-6,934 |
| Brielle | 16.9 | 0.10% | 0.09% | 17.7 | 14.07-24.09 | 3,578.5 | 1,490-7,519 |
| Bronckhorst | 35.7 | 0.21% | 0.18% | 16.3 | 12.90-21.21 | 2,780.5 | 1,290-5,128 |
| Brummen | 20.3 | 0.12% | 0.11% | 17.9 | 13.63-26.56 | 3,117.7 | 1,402-5,008 |
| Brunssum | 27.6 | 0.17% | 0.14% | 16.8 | 12.73-23.54 | 2,547.5 | 1,159-5,233 |
| Bunnik | 14.6 | 0.09% | 0.09% | 20.1 | 15.85-26.26 | 3,678.9 | 1,580-6,781 |
| Bunschoten | 21.3 | 0.13% | 0.13% | 19.4 | 13.72-26.29 | 3,545.8 | 1,250-6,108 |
| Buren | 26.0 | 0.16% | 0.16% | 20.5 | 16.55-26.14 | 3,629.0 | 1,563-7,088 |
| Capelle aan den IJssel | 66.4 | 0.40% | 0.43% | 21.2 | 18.22-24.89 | 4,212.4 | 1,923-7,007 |
| Castricum | 35.3 | 0.21% | 0.21% | 19.7 | 16.09-24.86 | 3,717.5 | 1,210-6,768 |
| Coevorden | 34.6 | 0.21% | 0.15% | 14.0 | 10.65-18.66 | 2,055.6 | 542-3,990 |
| Cranendonck | 19.9 | 0.12% | 0.11% | 18.4 | 13.89-24.54 | 3,245.8 | 1,350-6,297 |
| Cuijk | 24.3 | 0.15% | 0.15% | 19.7 | 14.88-25.05 | 3,769.5 | 1,608-7,060 |
| Culemborg | 27.9 | 0.17% | 0.17% | 20.1 | 16.28-25.57 | 3,977.1 | 1,784-6,371 |
| Dalfsen | 27.9 | 0.17% | 0.14% | 16.7 | 12.56-23.78 | 2,453.1 | 724-4,932 |
| Dantumadiel | 18.5 | 0.11% | 0.09% | 15.4 | 10.96-21.45 | 2,594.3 | 1,078-4,805 |
| De Bilt | 42.2 | 0.25% | 0.25% | 19.5 | 16.05-25.26 | 3,820.6 | 1,400-7,104 |
| De Fryske Marren | 51.0 | 0.31% | 0.27% | 17.2 | 13.16-23.55 | 2,748.1 | 1,002-5,298 |
| De Ronde Venen | 43.6 | 0.26% | 0.27% | 20.2 | 16.62-24.65 | 4,045.5 | 1,985-7,115 |
| De Wolden | 23.4 | 0.14% | 0.11% | 15.5 | 12.09-21.23 | 2,947.1 | 1,238-5,915 |
| Delft | 102.7 | 0.62% | 0.75% | 24.1 | 21.15-27.74 | 3,739.0 | 1,854-7,022 |
| Delfzijl | 24.2 | 0.15% | 0.09% | 12.4 | 8.54-19.30 | 1,920.3 | 537-4,300 |
| Den Helder | 55.1 | 0.33% | 0.30% | 18.2 | 15.05-22.16 | 3,128.2 | 1,276-5,776 |
| Deurne | 31.8 | 0.19% | 0.18% | 18.5 | 14.68-23.98 | 2,973.5 | 875-5,605 |
| Deventer | 99.6 | 0.60% | 0.54% | 17.8 | 14.71-22.18 | 2,632.1 | 901-4,974 |
| Diemen | 28.6 | 0.17% | 0.22% | 24.9 | 20.95-30.65 | 3,534.8 | 1,574-6,586 |
| Dinkelland | 25.9 | 0.16% | 0.12% | 14.7 | 10.85-21.57 | 2,562.4 | 785-5,175 |
| Doesburg | 10.5 | 0.06% | 0.06% | 18.4 | 13.56-25.22 | 3,094.5 | 1,232-5,683 |
| Doetinchem | 57.2 | 0.34% | 0.30% | 17.4 | 13.87-21.46 | 2,754.1 | 1,125-5,685 |
| Dongen | 25.5 | 0.15% | 0.15% | 19.2 | 15.64-25.85 | 3,624.7 | 1,357-6,324 |
| Dordrecht | 118.1 | 0.71% | 0.75% | 20.9 | 18.09-24.06 | 3,840.9 | 1,650-6,796 |
| Drechterland | 19.0 | 0.11% | 0.11% | 18.5 | 13.33-23.95 | 3,780.9 | 1,434-6,861 |
| Drimmelen | 26.5 | 0.16% | 0.15% | 18.9 | 15.34-24.26 | 3,558.5 | 1,499-5,830 |
| Dronten | 40.2 | 0.24% | 0.23% | 18.7 | 15.18-23.90 | 3,102.5 | 1,055-5,639 |
| Druten | 18.3 | 0.11% | 0.11% | 19.8 | 15.17-24.53 | 3,721.5 | 1,618-6,695 |
| Duiven | 24.9 | 0.15% | 0.14% | 18.6 | 14.05-24.78 | 3,418.0 | 1,207-6,255 |
| Echt-Susteren | 31.1 | 0.19% | 0.16% | 17.2 | 13.07-22.36 | 3,016.1 | 1,063-5,511 |
| Edam-Volendam | 35.7 | 0.21% | 0.21% | 19.3 | 15.13-26.03 | 3,279.2 | 1,327-5,684 |
| Ede | 115.2 | 0.69% | 0.70% | 20.0 | 17.22-24.15 | 3,669.9 | 1,462-6,849 |
| Eemnes | 8.6 | 0.05% | 0.05% | 20.4 | 15.08-27.88 | 4,000.8 | 1,747-6,591 |
| Eersel | 18.6 | 0.11% | 0.11% | 19.6 | 15.34-25.65 | 3,690.7 | 1,737-7,108 |
| Eijsden-Margraten | 25.1 | 0.15% | 0.13% | 17.1 | 12.47-24.20 | 2,562.5 | 886-4,976 |
| Eindhoven | 230.5 | 1.38% | 1.54% | 22.0 | 18.54-24.95 | 3,224.8 | 1,186-6,081 |
| Elburg | 22.5 | 0.14% | 0.13% | 19.0 | 14.47-25.72 | 3,224.6 | 1,271-6,218 |
| Emmen | 106.4 | 0.64% | 0.48% | 14.8 | 11.88-19.48 | 2,033.0 | 435-4,570 |
| Enkhuizen | 18.0 | 0.11% | 0.11% | 19.4 | 14.63-25.21 | 3,740.7 | 1,771-6,506 |
| Enschede | 158.2 | 0.95% | 0.76% | 15.9 | 13.49-19.15 | 2,187.1 | 597-5,108 |
| Epe | 32.8 | 0.20% | 0.18% | 18.2 | 13.83-23.74 | 2,692.7 | 1,107-5,393 |
| Ermelo | 26.3 | 0.16% | 0.15% | 19.4 | 15.28-24.58 | 3,711.5 | 1,785-6,302 |
| Etten-Leur | 43.2 | 0.26% | 0.26% | 19.5 | 15.26-24.71 | 3,380.9 | 1,284-5,941 |
| Geertruidenberg | 21.0 | 0.13% | 0.13% | 19.8 | 15.98-24.22 | 3,822.7 | 1,588-6,969 |
| Geldrop-Mierlo | 39.1 | 0.23% | 0.22% | 18.9 | 15.08-26.31 | 3,927.1 | 1,420-7,249 |
| Gemert-Bakel | 30.0 | 0.18% | 0.17% | 18.8 | 14.99-25.31 | 2,719.4 | 876-5,010 |
| Gennep | 16.7 | 0.10% | 0.09% | 17.6 | 13.64-23.44 | 3,368.1 | 1,689-6,461 |
| Gilze en Rijen | 25.9 | 0.16% | 0.15% | 19.1 | 14.34-27.39 | 3,154.7 | 1,180-6,552 |
| Goeree-Overflakkee | 49.2 | 0.30% | 0.26% | 17.4 | 13.43-23.99 | 3,127.5 | 1,276-5,724 |
| Goes | 37.1 | 0.22% | 0.18% | 15.6 | 11.32-20.68 | 2,432.0 | 812-5,096 |
| Goirle | 23.1 | 0.14% | 0.14% | 19.3 | 15.46-23.94 | 3,897.9 | 1,650-6,450 |
| Gooise Meren | 57.1 | 0.34% | 0.37% | 21.1 | 17.77-26.49 | 4,051.9 | 1,960-7,028 |
| Gorinchem | 35.9 | 0.22% | 0.23% | 20.8 | 17.08-27.62 | 3,878.7 | 1,680-6,685 |
| Gouda | 72.6 | 0.44% | 0.46% | 20.7 | 17.51-25.16 | 3,499.1 | 1,524-5,791 |
| Grave | 12.1 | 0.07% | 0.07% | 20.1 | 14.48-28.37 | 3,410.6 | 1,649-6,048 |
| Groningen | 203.2 | 1.22% | 1.08% | 17.5 | 14.41-21.50 | 2,364.9 | 616-4,509 |

Continued on next page

| Municipality | Population size (×1000) | Population percentage | Percentage of transmissions | Transmission risk | (90% range) | Seed Risk | (90% range) |
|---|---|---|---|---|---|---|---|
| Gulpen-Wittem | 13.8 | 0.08% | 0.07% | 16.1 | 10.95-22.04 | 2,736.2 | 1,329-4,369 |
| Haaksbergen | 23.6 | 0.14% | 0.11% | 15.7 | 12.06-21.45 | 2,624.0 | 970-4,710 |
| Haaren | 13.6 | 0.08% | 0.08% | 19.8 | 14.97-28.45 | 3,348.0 | 1,368-5,476 |
| Haarlem | 160.5 | 0.96% | 1.08% | 22.2 | 19.68-25.78 | 3,944.2 | 1,885-6,788 |
| Haarlemmermeer | 147.3 | 0.88% | 1.01% | 22.5 | 19.22-25.87 | 4,054.8 | 2,240-6,909 |
| Halderberge | 29.8 | 0.18% | 0.16% | 17.6 | 13.81-22.46 | 3,254.0 | 1,206-7,346 |
| Hardenberg | 60.3 | 0.36% | 0.31% | 17.0 | 14.22-23.36 | 2,781.4 | 776-6,133 |
| Harderwijk | 46.7 | 0.28% | 0.27% | 18.9 | 14.62-23.83 | 3,178.9 | 1,268-5,663 |
| Hardinxveld-Giessendam | 17.5 | 0.11% | 0.10% | 19.7 | 14.58-30.24 | 3,820.5 | 1,449-7,157 |
| Harlingen | 15.3 | 0.09% | 0.07% | 16.0 | 11.49-21.64 | 2,673.5 | 1,084-5,422 |
| Hattem | 11.6 | 0.07% | 0.06% | 18.1 | 13.14-25.91 | 3,074.2 | 1,253-5,556 |
| Heemskerk | 38.8 | 0.23% | 0.26% | 21.8 | 17.73-27.39 | 4,283.8 | 1,703-7,469 |
| Heemstede | 26.8 | 0.16% | 0.17% | 20.5 | 16.63-25.34 | 4,147.3 | 1,777-7,788 |
| Heerde | 18.1 | 0.11% | 0.10% | 17.6 | 13.21-22.59 | 3,016.9 | 1,527-5,096 |
| Heerenveen | 49.6 | 0.30% | 0.25% | 16.8 | 12.87-21.13 | 2,629.6 | 871-5,465 |
| Heerhugowaard | 56.0 | 0.34% | 0.32% | 18.6 | 14.88-22.94 | 3,278.5 | 1,202-6,401 |
| Heerlen | 86.6 | 0.52% | 0.43% | 16.3 | 13.80-19.40 | 2,803.6 | 758-5,466 |
| Heeze-Leende | 15.5 | 0.09% | 0.09% | 18.2 | 13.81-22.81 | 3,183.3 | 1,434-5,307 |
| Heiloo | 23.0 | 0.14% | 0.13% | 18.8 | 15.12-23.42 | 3,578.6 | 1,461-6,006 |
| Hellendoorn | 35.4 | 0.21% | 0.18% | 16.6 | 12.61-21.65 | 2,501.6 | 1,180-4,460 |
| Hellevoetsluis | 39.6 | 0.24% | 0.23% | 18.9 | 14.26-26.80 | 3,235.8 | 1,317-5,701 |
| Helmond | 90.9 | 0.55% | 0.53% | 19.1 | 15.81-23.36 | 2,962.5 | 965-6,441 |
| Hendrik-Ido-Ambacht | 30.4 | 0.18% | 0.18% | 19.2 | 15.67-23.16 | 3,667.4 | 1,608-7,296 |
| Hengelo | 80.3 | 0.48% | 0.40% | 16.4 | 12.74-21.67 | 2,152.7 | 598-4,737 |
| Het Hogeland | 11.2 | 0.07% | 0.05% | 14.9 | 9.79-27.54 | 2,155.4 | 0-5,351 |
| Heumen | 16.0 | 0.10% | 0.09% | 17.8 | 13.25-24.91 | 3,335.6 | 1,253-5,694 |
| Heusden | 43.6 | 0.26% | 0.27% | 20.3 | 16.09-27.36 | 3,856.2 | 1,883-6,956 |
| Hillegom | 21.4 | 0.13% | 0.13% | 20.0 | 15.60-25.54 | 4,145.5 | 2,011-7,112 |
| Hilvarenbeek | 14.9 | 0.09% | 0.12% | 26.4 | 20.79-35.50 | 3,391.7 | 1,159-6,304 |
| Hilversum | 89.7 | 0.54% | 0.61% | 22.4 | 19.37-27.52 | 3,854.9 | 1,747-8,141 |
| Hoeksche Waard | 11.2 | 0.07% | 0.06% | 18.4 | 12.99-26.26 | 3,579.0 | 1,584-7,229 |
| Hof van Twente | 34.5 | 0.21% | 0.17% | 16.1 | 12.61-19.79 | 2,408.0 | 960-4,567 |
| Hollands Kroon | 46.9 | 0.28% | 0.25% | 17.5 | 12.86-23.01 | 2,633.1 | 781-4,967 |
| Hoogeveen | 55.0 | 0.33% | 0.27% | 16.1 | 12.41-21.15 | 2,367.3 | 730-4,771 |
| Hoorn | 72.6 | 0.44% | 0.43% | 19.4 | 15.90-24.39 | 3,295.1 | 1,415-5,624 |
| Horst aan de Maas | 41.7 | 0.25% | 0.22% | 17.8 | 13.97-22.01 | 3,035.9 | 1,312-5,588 |
| Houten | 49.5 | 0.30% | 0.30% | 20.2 | 16.51-25.17 | 3,774.2 | 1,431-6,108 |
| Huizen | 40.9 | 0.25% | 0.25% | 19.9 | 15.97-24.76 | 3,771.4 | 1,914-7,230 |
| Hulst | 27.0 | 0.16% | 0.13% | 16.3 | 12.56-21.93 | 2,767.6 | 823-4,966 |
| IJsselstein | 33.6 | 0.20% | 0.22% | 21.7 | 17.91-28.20 | 3,611.1 | 1,565-6,196 |
| Kaag en Braassem | 26.4 | 0.16% | 0.17% | 20.6 | 16.07-27.04 | 3,550.3 | 1,437-6,493 |
| Kampen | 53.2 | 0.32% | 0.28% | 17.5 | 14.66-21.67 | 3,134.2 | 1,067-5,934 |
| Kapelle | 12.4 | 0.07% | 0.06% | 15.2 | 10.41-23.96 | 2,471.7 | 914-4,790 |
| Katwijk | 64.6 | 0.39% | 0.44% | 22.2 | 18.15-27.27 | 3,941.2 | 1,830-6,245 |
| Kerkrade | 45.1 | 0.27% | 0.21% | 15.3 | 12.42-19.72 | 2,795.9 | 776-5,517 |
| Koggenland | 22.4 | 0.13% | 0.13% | 18.4 | 14.68-23.80 | 3,381.4 | 1,407-6,745 |
| Krimpen aan den IJssel | 28.8 | 0.17% | 0.19% | 22.0 | 18.03-26.55 | 4,916.2 | 2,442-8,317 |
| Krimpenerwaard | 55.5 | 0.33% | 0.33% | 19.8 | 16.26-24.97 | 3,810.6 | 1,768-6,347 |
| Laarbeek | 21.8 | 0.13% | 0.14% | 20.7 | 15.33-27.79 | 3,485.1 | 1,445-6,649 |
| Landerd | 14.8 | 0.09% | 0.09% | 19.2 | 14.52-27.09 | 3,593.2 | 1,376-6,024 |
| Landgraaf | 37.0 | 0.22% | 0.17% | 15.4 | 12.26-19.63 | 2,626.3 | 755-4,926 |
| Landsmeer | 11.0 | 0.07% | 0.08% | 22.5 | 17.17-29.25 | 4,079.8 | 1,486-6,992 |
| Langedijk | 27.5 | 0.17% | 0.15% | 17.8 | 14.55-24.23 | 3,120.6 | 1,304-5,496 |
| Lansingerland | 61.1 | 0.37% | 0.39% | 21.1 | 17.35-25.38 | 4,135.4 | 1,935-7,039 |
| Laren | 10.7 | 0.06% | 0.07% | 20.4 | 15.15-25.77 | 4,038.0 | 1,844-7,327 |
| Leeuwarden | 122.4 | 0.73% | 0.65% | 17.4 | 14.28-21.83 | 2,491.5 | 1,091-5,253 |
| Leiden | 124.1 | 0.75% | 0.88% | 23.3 | 20.26-26.90 | 4,008.6 | 1,582-7,106 |
| Leiderdorp | 26.7 | 0.16% | 0.16% | 20.0 | 16.03-26.36 | 3,909.1 | 1,726-7,186 |
| Leidschendam-Voorburg | 74.8 | 0.45% | 0.49% | 21.7 | 19.23-25.89 | 4,242.6 | 2,144-7,494 |
| Lelystad | 77.6 | 0.47% | 0.44% | 18.8 | 15.63-23.49 | 3,159.0 | 1,226-6,243 |
| Leudal | 35.3 | 0.21% | 0.18% | 16.8 | 12.95-22.43 | 2,748.6 | 820-5,110 |
| Leusden | 29.2 | 0.18% | 0.18% | 20.0 | 15.21-27.34 | 4,330.3 | 2,162-8,242 |

Continued on next page

| Municipality | Population size (×1000) | Population percentage | Percentage of transmissions | Transmission risk | (90% range) | Seed Risk | (90% range) |
|---|---|---|---|---|---|---|---|
| Lingewaard | 45.9 | 0.28% | 0.27% | 19.1 | 15.11-25.30 | 3,246.2 | 1,373-5,654 |
| Lisse | 22.4 | 0.13% | 0.14% | 20.6 | 16.34-25.90 | 3,956.7 | 1,817-7,530 |
| Lochem | 33.1 | 0.20% | 0.16% | 16.3 | 12.47-22.73 | 2,610.8 | 1,144-4,999 |
| Loon op Zand | 22.6 | 0.14% | 0.14% | 20.1 | 16.15-24.81 | 3,998.0 | 1,863-6,974 |
| Lopik | 13.9 | 0.08% | 0.09% | 20.9 | 15.91-28.09 | 3,409.0 | 1,371-6,558 |
| Loppersum | 9.0 | 0.05% | 0.04% | 13.4 | 8.80-22.06 | 2,390.2 | 653-5,071 |
| Losser | 22.2 | 0.13% | 0.09% | 13.4 | 9.16-20.30 | 2,049.8 | 434-4,989 |
| Maasdriel | 24.0 | 0.14% | 0.15% | 20.2 | 14.82-27.29 | 3,337.1 | 1,507-5,771 |
| Maasgouw | 23.2 | 0.14% | 0.12% | 17.5 | 13.22-22.15 | 2,529.7 | 1,006-4,341 |
| Maassluis | 32.2 | 0.19% | 0.21% | 21.1 | 17.04-25.86 | 3,563.7 | 1,364-6,126 |
| Maastricht | 121.7 | 0.73% | 0.64% | 17.2 | 14.40-20.56 | 2,419.7 | 900-4,495 |
| Medemblik | 44.2 | 0.27% | 0.23% | 17.2 | 14.22-22.67 | 3,209.6 | 1,176-6,106 |
| Meerssen | 18.5 | 0.11% | 0.09% | 16.7 | 12.24-24.51 | 3,013.6 | 1,014-5,726 |
| Meierijstad | 80.3 | 0.48% | 0.50% | 20.5 | 17.28-25.40 | 3,665.9 | 1,805-6,336 |
| Meppel | 33.0 | 0.20% | 0.16% | 16.4 | 13.10-21.13 | 2,770.4 | 1,078-5,205 |
| Middelburg | 48.3 | 0.29% | 0.21% | 14.1 | 11.28-17.62 | 2,237.8 | 600-5,012 |
| Midden-Delfland | 19.1 | 0.11% | 0.15% | 25.4 | 20.43-33.06 | 3,855.5 | 1,659-6,096 |
| Midden-Drenthe | 32.5 | 0.20% | 0.16% | 16.2 | 12.11-26.40 | 2,181.0 | 610-4,372 |
| Midden-Groningen | 60.5 | 0.36% | 0.26% | 14.4 | 11.15-18.72 | 2,346.1 | 399-4,954 |
| Mill en Sint Hubert | 10.5 | 0.06% | 0.06% | 20.3 | 14.93-27.52 | 3,336.6 | 1,414-5,689 |
| Moerdijk | 36.6 | 0.22% | 0.21% | 19.1 | 15.40-23.47 | 3,580.6 | 1,532-6,288 |
| Molenlanden | 11.2 | 0.07% | 0.07% | 21.2 | 15.88-28.54 | 4,194.1 | 1,500-8,430 |
| Montferland | 35.3 | 0.21% | 0.18% | 17.3 | 13.37-22.18 | 2,725.9 | 865-4,914 |
| Montfoort | 13.4 | 0.08% | 0.09% | 21.1 | 15.50-26.80 | 3,797.7 | 1,646-6,809 |
| Mook en Middelaar | 7.3 | 0.04% | 0.04% | 18.0 | 13.39-23.25 | 3,247.6 | 1,444-5,968 |
| Neder-Betuwe | 23.3 | 0.14% | 0.14% | 19.4 | 15.21-24.97 | 3,731.2 | 1,265-7,131 |
| Nederweert | 16.6 | 0.10% | 0.09% | 18.6 | 14.30-25.56 | 3,149.0 | 1,043-5,551 |
| Nieuwegein | 62.5 | 0.38% | 0.41% | 21.8 | 19.19-25.52 | 4,335.7 | 2,222-7,406 |
| Nieuwkoop | 28.0 | 0.17% | 0.18% | 20.7 | 16.57-26.61 | 3,735.2 | 1,445-6,804 |
| Nijkerk | 42.3 | 0.25% | 0.25% | 19.7 | 15.41-24.49 | 3,395.7 | 1,636-6,017 |
| Nijmegen | 176.6 | 1.06% | 1.13% | 21.0 | 18.20-25.08 | 3,216.9 | 1,225-5,595 |
| Nissewaard | 84.2 | 0.51% | 0.51% | 20.0 | 17.04-23.94 | 3,302.6 | 1,506-5,739 |
| Noardeast-FryslÃ¢n | 11.2 | 0.07% | 0.04% | 13.1 | 9.21-19.42 | 2,148.7 | 475-4,140 |
| Noord-Beveland | 6.7 | 0.04% | 0.03% | 14.4 | 9.35-21.68 | 2,336.6 | 553-5,230 |
| Noordenveld | 30.8 | 0.18% | 0.14% | 14.7 | 11.28-18.98 | 2,300.1 | 775-4,309 |
| Noordoostpolder | 46.3 | 0.28% | 0.25% | 17.6 | 13.98-22.90 | 2,532.0 | 797-4,811 |
| Noordwijk | 25.8 | 0.15% | 0.16% | 19.9 | 15.14-27.46 | 3,800.8 | 1,632-6,674 |
| Nuenen, Gerwen en Nederwetten | 22.7 | 0.14% | 0.13% | 18.2 | 14.22-24.87 | 3,379.5 | 1,188-6,615 |
| Nunspeet | 27.1 | 0.16% | 0.15% | 18.3 | 13.81-26.52 | 3,422.9 | 1,371-6,349 |
| Oegstgeest | 23.9 | 0.14% | 0.17% | 23.8 | 18.96-30.66 | 4,005.2 | 1,904-7,004 |
| Oirschot | 18.1 | 0.11% | 0.11% | 19.8 | 14.51-26.08 | 3,840.0 | 1,506-7,185 |
| Oisterwijk | 25.4 | 0.15% | 0.15% | 18.9 | 14.30-27.52 | 3,316.9 | 966-5,904 |
| Oldambt | 37.5 | 0.23% | 0.16% | 13.6 | 9.96-20.88 | 1,591.1 | 390-3,538 |
| Oldebroek | 23.1 | 0.14% | 0.13% | 17.9 | 13.24-25.06 | 3,041.1 | 1,110-6,218 |
| Oldenzaal | 31.1 | 0.19% | 0.13% | 14.2 | 10.72-17.74 | 2,391.4 | 787-4,719 |
| Olst-Wijhe | 17.6 | 0.11% | 0.10% | 17.8 | 13.29-22.11 | 3,268.6 | 1,076-5,801 |
| Ommen | 17.4 | 0.10% | 0.09% | 17.2 | 10.88-27.00 | 2,843.2 | 861-5,243 |
| Oost Gelre | 29.2 | 0.18% | 0.14% | 15.8 | 11.97-20.22 | 2,405.2 | 610-5,513 |
| Oosterhout | 54.7 | 0.33% | 0.34% | 20.3 | 17.26-25.67 | 3,891.7 | 1,467-7,088 |
| Ooststellingwerf | 24.9 | 0.15% | 0.12% | 16.2 | 12.09-24.40 | 2,465.0 | 965-4,447 |
| Oostzaan | 9.2 | 0.06% | 0.06% | 21.7 | 14.84-28.42 | 4,005.3 | 2,012-7,014 |
| Opmeer | 11.2 | 0.07% | 0.07% | 19.2 | 13.94-31.36 | 3,798.7 | 1,506-6,373 |
| Opsterland | 29.4 | 0.18% | 0.14% | 15.7 | 11.94-22.44 | 2,628.8 | 809-4,989 |
| Oss | 91.0 | 0.55% | 0.55% | 20.1 | 17.17-23.38 | 3,652.8 | 1,494-7,285 |
| Oude IJsselstreek | 38.8 | 0.23% | 0.19% | 16.1 | 12.33-21.29 | 2,657.4 | 629-4,892 |
| Ouder-Amstel | 13.3 | 0.08% | 0.09% | 23.0 | 17.78-30.69 | 4,350.8 | 1,734-7,459 |
| Oudewater | 9.7 | 0.06% | 0.07% | 22.5 | 16.79-31.65 | 4,176.1 | 2,118-6,961 |
| Overbetuwe | 47.0 | 0.28% | 0.27% | 18.6 | 15.86-22.10 | 3,578.2 | 1,380-6,855 |
| Papendrecht | 31.8 | 0.19% | 0.19% | 19.3 | 15.94-23.88 | 3,893.4 | 1,663-7,281 |
| Peel en Maas | 42.9 | 0.26% | 0.23% | 17.5 | 13.67-24.05 | 2,859.5 | 647-5,176 |
| Pekela | 11.9 | 0.07% | 0.05% | 13.5 | 8.83-18.99 | 1,970.8 | 631-3,980 |

Continued on next page

| Municipality | Population size (×1000) | Population percentage | Percentage of transmissions | Transmission risk | (90% range) | Seed Risk | (90% range) |
|---|---|---|---|---|---|---|---|
| Pijnacker-Nootdorp | 53.6 | 0.32% | 0.41% | 25.2 | 20.72-31.35 | 4,370.6 | 1,574-7,835 |
| Purmerend | 79.6 | 0.48% | 0.50% | 20.8 | 17.12-25.90 | 3,581.6 | 1,228-7,087 |
| Putten | 23.8 | 0.14% | 0.14% | 19.9 | 16.72-23.98 | 3,865.9 | 1,544-7,209 |
| Raalte | 36.9 | 0.22% | 0.19% | 16.6 | 12.71-22.41 | 2,972.4 | 943-5,860 |
| Reimerswaal | 22.2 | 0.13% | 0.11% | 16.0 | 12.37-20.96 | 2,696.5 | 885-5,733 |
| Renkum | 30.9 | 0.19% | 0.18% | 19.5 | 15.48-24.32 | 3,738.5 | 1,243-7,054 |
| Renswoude | 4.6 | 0.03% | 0.03% | 25.0 | 14.70-41.05 | 3,865.3 | 1,819-6,089 |
| Reusel-De Mierden | 12.6 | 0.08% | 0.08% | 19.7 | 13.95-28.87 | 3,299.0 | 1,431-6,293 |
| Rheden | 43.3 | 0.26% | 0.23% | 17.5 | 14.31-22.34 | 2,707.8 | 913-5,149 |
| Rhenen | 19.4 | 0.12% | 0.12% | 21.2 | 17.38-27.38 | 3,889.0 | 1,847-7,461 |
| Ridderkerk | 45.7 | 0.27% | 0.27% | 19.7 | 16.08-24.58 | 3,498.3 | 1,515-6,546 |
| Rijssen-Holten | 37.8 | 0.23% | 0.19% | 16.8 | 12.92-21.83 | 2,548.6 | 686-4,865 |
| Rijswijk | 52.7 | 0.32% | 0.36% | 22.4 | 19.42-26.90 | 3,827.6 | 1,982-6,068 |
| Roerdalen | 20.1 | 0.12% | 0.10% | 15.9 | 12.64-21.02 | 3,070.8 | 1,182-5,787 |
| Roermond | 57.4 | 0.34% | 0.30% | 17.3 | 14.10-21.74 | 3,022.8 | 1,227-5,725 |
| Roosendaal | 76.6 | 0.46% | 0.44% | 19.1 | 16.27-22.89 | 3,286.4 | 1,191-6,042 |
| Rotterdam | 641.3 | 3.85% | 4.65% | 23.9 | 21.88-25.99 | 3,791.2 | 1,819-6,356 |
| Rozendaal | 1.3 | 0.01% | 0.00% | 12.2 | 6.19-23.94 | 3,393.8 | 1,324-5,438 |
| Rucphen | 22.2 | 0.13% | 0.13% | 19.2 | 14.37-26.18 | 3,455.0 | 1,391-6,238 |
| Schagen | 45.9 | 0.28% | 0.27% | 19.2 | 15.05-26.13 | 3,979.7 | 1,622-6,865 |
| Scherpenzeel | 9.5 | 0.06% | 0.06% | 19.6 | 14.03-26.35 | 3,334.8 | 1,374-6,117 |
| Schiedam | 77.4 | 0.46% | 0.61% | 25.8 | 22.48-29.87 | 3,793.7 | 1,447-6,614 |
| Schiermonnikoog | 0.5 | 0.00% | 0.00% | 8.6 | 1.76-18.69 | 3,262.6 | 1,346-6,054 |
| Schouwen-Duiveland | 33.3 | 0.20% | 0.16% | 15.5 | 12.05-21.61 | 2,800.2 | 823-6,367 |
| Simpelveld | 9.9 | 0.06% | 0.05% | 15.6 | 10.40-23.35 | 2,592.5 | 917-5,042 |
| Sint Anthonis | 11.2 | 0.07% | 0.06% | 18.2 | 13.73-24.54 | 3,386.0 | 1,202-6,666 |
| Sint-Michielsgestel | 28.5 | 0.17% | 0.18% | 20.9 | 16.91-26.02 | 3,724.1 | 1,417-6,198 |
| Sittard-Geleen | 92.2 | 0.55% | 0.47% | 17.0 | 14.17-20.92 | 2,660.5 | 717-5,597 |
| Sliedrecht | 24.6 | 0.15% | 0.15% | 20.6 | 15.90-27.38 | 3,820.0 | 1,880-6,867 |
| Sluis | 23.0 | 0.14% | 0.09% | 12.9 | 9.49-18.89 | 2,409.4 | 533-5,018 |
| Smallingerland | 55.4 | 0.33% | 0.28% | 16.7 | 12.73-22.52 | 2,420.4 | 771-4,827 |
| Soest | 45.6 | 0.27% | 0.27% | 19.6 | 15.87-23.30 | 3,997.9 | 1,567-6,967 |
| Someren | 18.8 | 0.11% | 0.11% | 19.2 | 14.18-25.48 | 2,728.4 | 938-4,666 |
| Son en Breugel | 16.2 | 0.10% | 0.10% | 20.2 | 15.71-26.88 | 3,423.1 | 1,619-6,028 |
| Stadskanaal | 31.4 | 0.19% | 0.14% | 14.3 | 9.74-22.10 | 2,073.9 | 414-4,474 |
| Staphorst | 16.4 | 0.10% | 0.09% | 17.1 | 12.37-23.64 | 2,364.4 | 979-4,892 |
| Stede Broec | 21.3 | 0.13% | 0.12% | 18.6 | 14.16-23.72 | 2,957.1 | 977-5,751 |
| Steenbergen | 25.0 | 0.15% | 0.14% | 18.4 | 13.59-25.95 | 2,391.1 | 924-4,040 |
| Steenwijkerland | 43.3 | 0.26% | 0.23% | 17.2 | 13.72-22.28 | 2,916.4 | 1,305-5,721 |
| Stein | 24.4 | 0.15% | 0.13% | 16.9 | 12.56-23.77 | 2,923.0 | 1,103-4,861 |
| Stichtse Vecht | 63.9 | 0.38% | 0.39% | 20.2 | 17.03-23.91 | 3,556.6 | 1,290-6,148 |
| SÃºdwest-FryslÃ¢n | 89.0 | 0.53% | 0.46% | 16.9 | 13.73-20.95 | 2,764.5 | 679-5,896 |
| Terneuzen | 54.1 | 0.32% | 0.25% | 15.1 | 11.16-20.38 | 2,563.3 | 834-5,339 |
| Terschelling | 4.5 | 0.03% | 0.02% | 15.4 | 7.80-27.96 | 2,376.2 | 764-4,760 |
| Texel | 13.0 | 0.08% | 0.07% | 16.6 | 11.81-23.38 | 2,902.0 | 1,005-6,262 |
| Teylingen | 36.5 | 0.22% | 0.22% | 19.9 | 16.38-25.25 | 4,023.2 | 2,095-6,619 |
| Tholen | 25.2 | 0.15% | 0.14% | 18.0 | 13.31-24.96 | 2,995.5 | 933-6,290 |
| Tiel | 41.5 | 0.25% | 0.24% | 19.1 | 15.81-22.99 | 3,726.5 | 1,582-6,936 |
| Tilburg | 216.5 | 1.30% | 1.38% | 21.0 | 18.44-24.10 | 3,676.9 | 1,639-6,851 |
| Tubbergen | 20.6 | 0.12% | 0.10% | 16.2 | 10.97-21.33 | 2,510.7 | 980-5,115 |
| Twenterand | 33.6 | 0.20% | 0.16% | 15.6 | 12.14-20.80 | 2,553.3 | 792-4,586 |
| Tynaarlo | 33.2 | 0.20% | 0.14% | 14.1 | 10.22-18.88 | 1,902.0 | 608-3,855 |
| Tytsjerksteradiel | 31.3 | 0.19% | 0.15% | 16.0 | 12.35-19.74 | 2,711.2 | 1,054-5,071 |
| Uden | 41.2 | 0.25% | 0.24% | 19.0 | 15.16-23.90 | 3,101.4 | 957-5,337 |
| Uitgeest | 13.0 | 0.08% | 0.09% | 21.9 | 15.98-30.23 | 3,565.2 | 1,249-7,122 |
| Uithoorn | 28.9 | 0.17% | 0.18% | 20.6 | 16.50-26.19 | 3,864.8 | 1,622-6,826 |
| Urk | 20.2 | 0.12% | 0.12% | 19.3 | 14.69-28.76 | 2,950.2 | 939-6,038 |
| Utrecht | 350.4 | 2.10% | 2.53% | 23.8 | 21.26-26.93 | 3,516.2 | 1,644-6,463 |
| Utrechtse Heuvelrug | 49.0 | 0.29% | 0.28% | 18.9 | 15.85-22.83 | 3,303.5 | 1,186-6,398 |
| Vaals | 9.6 | 0.06% | 0.05% | 16.8 | 11.44-23.88 | 2,845.0 | 1,039-5,466 |

Continued on next page

| Municipality | Population size (×1000) | Population percentage | Percentage of transmissions | Transmission risk | (90% range) | Seed Risk | (90% range) |
|---|---|---|---|---|---|---|---|
| Valkenburg aan de Geul | 16.0 | 0.10% | 0.08% | 15.9 | 10.79-21.88 | 2,844.7 | 1,010-5,497 |
| Valkenswaard | 30.4 | 0.18% | 0.17% | 18.5 | 15.02-22.85 | 3,398.5 | 1,250-5,832 |
| Veendam | 26.9 | 0.16% | 0.12% | 14.5 | 10.20-19.10 | 2,213.9 | 504-4,762 |
| Veenendaal | 64.9 | 0.39% | 0.38% | 19.5 | 16.90-23.66 | 3,717.5 | 1,676-7,590 |
| Veere | 21.2 | 0.13% | 0.09% | 13.8 | 9.52-21.77 | 2,024.6 | 544-4,211 |
| Veldhoven | 44.6 | 0.27% | 0.25% | 18.3 | 15.59-22.11 | 2,939.2 | 1,417-5,521 |
| Velsen | 67.7 | 0.41% | 0.42% | 20.4 | 17.17-23.69 | 3,715.1 | 1,244-6,520 |
| Venlo | 101.0 | 0.61% | 0.53% | 17.3 | 14.11-21.19 | 2,509.4 | 887-4,794 |
| Venray | 42.8 | 0.26% | 0.22% | 16.9 | 13.71-21.38 | 2,760.1 | 560-5,254 |
| Vijfheerenlanden | 11.2 | 0.07% | 0.07% | 21.5 | 16.53-29.18 | 4,327.9 | 1,720-7,993 |
| Vlaardingen | 71.8 | 0.43% | 0.53% | 24.4 | 20.69-28.71 | 3,275.3 | 1,204-5,792 |
| Vlieland | 0.6 | 0.00% | 0.00% | 10.2 | 3.11-20.76 | 3,195.0 | 1,163-5,883 |
| Vlissingen | 44.1 | 0.26% | 0.19% | 14.4 | 10.82-19.91 | 1,874.5 | 547-4,436 |
| Voerendaal | 12.0 | 0.07% | 0.06% | 16.1 | 12.00-21.59 | 2,972.0 | 827-5,493 |
| Voorschoten | 25.0 | 0.15% | 0.15% | 20.3 | 15.62-26.77 | 3,741.1 | 1,861-6,161 |
| Voorst | 23.9 | 0.14% | 0.13% | 18.0 | 13.57-23.11 | 2,932.5 | 1,122-5,460 |
| Vught | 26.0 | 0.16% | 0.16% | 19.7 | 16.18-25.20 | 3,863.2 | 1,857-6,477 |
| Waadhoeke | 45.6 | 0.27% | 0.23% | 16.8 | 13.46-21.21 | 2,630.4 | 885-5,092 |
| Waalre | 16.9 | 0.10% | 0.10% | 19.5 | 14.87-26.09 | 3,754.9 | 1,739-6,921 |
| Waalwijk | 47.7 | 0.29% | 0.27% | 18.9 | 15.11-22.58 | 3,340.3 | 1,106-6,333 |
| Waddinxveen | 27.9 | 0.17% | 0.17% | 20.6 | 16.17-25.58 | 3,930.9 | 1,939-7,261 |
| Wageningen | 38.1 | 0.23% | 0.24% | 20.6 | 17.30-24.60 | 3,454.7 | 1,368-6,051 |
| Wassenaar | 25.6 | 0.15% | 0.20% | 26.0 | 21.96-31.25 | 3,424.8 | 1,686-5,679 |
| Waterland | 16.8 | 0.10% | 0.11% | 20.8 | 16.46-26.29 | 3,764.0 | 1,235-6,234 |
| Weert | 49.1 | 0.29% | 0.25% | 16.8 | 13.85-20.69 | 2,500.5 | 888-4,503 |
| Weesp | 18.8 | 0.11% | 0.12% | 21.9 | 17.43-28.83 | 3,654.3 | 1,877-6,289 |
| West Betuwe | 11.2 | 0.07% | 0.07% | 20.8 | 15.93-27.61 | 3,770.7 | 1,270-6,817 |
| West Maas en Waal | 18.5 | 0.11% | 0.11% | 19.2 | 15.57-27.83 | 3,386.2 | 1,378-5,720 |
| Westerkwartier | 11.2 | 0.07% | 0.05% | 14.3 | 8.53-23.76 | 1,736.5 | 592-2,943 |
| Westerveld | 18.9 | 0.11% | 0.09% | 14.8 | 11.09-19.77 | 2,368.1 | 800-4,240 |
| Westervoort | 14.5 | 0.09% | 0.08% | 18.7 | 14.28-25.08 | 3,231.7 | 1,540-5,992 |
| Westerwolde | 24.8 | 0.15% | 0.10% | 13.1 | 9.10-20.59 | 1,784.2 | 348-3,986 |
| Westland | 108.3 | 0.65% | 0.66% | 20.0 | 17.55-23.92 | 3,773.5 | 1,648-7,076 |
| Weststellingwerf | 25.4 | 0.15% | 0.13% | 16.6 | 12.47-21.98 | 2,785.0 | 787-5,216 |
| Westvoorne | 14.3 | 0.09% | 0.07% | 17.0 | 12.58-23.12 | 2,706.8 | 914-5,167 |
| Wierden | 23.9 | 0.14% | 0.13% | 17.3 | 12.22-25.29 | 2,631.5 | 1,017-4,843 |
| Wijchen | 40.4 | 0.24% | 0.24% | 19.7 | 16.42-23.17 | 3,794.6 | 1,724-6,855 |
| Wijdemeren | 23.4 | 0.14% | 0.15% | 21.1 | 17.50-29.10 | 4,011.7 | 1,796-6,742 |
| Wijk bij Duurstede | 23.3 | 0.14% | 0.14% | 20.1 | 15.67-26.41 | 3,756.8 | 1,650-6,654 |
| Winterswijk | 28.5 | 0.17% | 0.14% | 16.1 | 11.59-24.07 | 2,797.0 | 745-5,631 |
| Woensdrecht | 21.4 | 0.13% | 0.12% | 18.1 | 14.02-25.27 | 3,297.7 | 1,073-5,457 |
| Woerden | 51.5 | 0.31% | 0.31% | 20.1 | 16.85-23.81 | 3,467.1 | 1,520-6,043 |
| Wormerland | 15.8 | 0.09% | 0.11% | 22.0 | 16.82-30.29 | 4,305.3 | 2,191-7,022 |
| Woudenberg | 12.6 | 0.08% | 0.08% | 21.5 | 15.57-32.67 | 3,639.9 | 1,557-6,633 |
| Zaanstad | 155.1 | 0.93% | 1.06% | 22.5 | 19.73-26.76 | 3,815.9 | 1,666-6,333 |
| Zaltbommel | 27.9 | 0.17% | 0.17% | 19.6 | 15.83-25.43 | 3,792.0 | 1,420-7,160 |
| Zandvoort | 16.3 | 0.10% | 0.10% | 19.8 | 15.23-24.60 | 3,501.7 | 1,573-5,844 |
| Zeewolde | 21.8 | 0.13% | 0.14% | 20.7 | 16.04-29.57 | 3,542.9 | 1,542-6,280 |
| Zeist | 63.6 | 0.38% | 0.38% | 19.7 | 16.72-22.93 | 3,528.3 | 1,682-6,782 |
| Zevenaar | 43.0 | 0.26% | 0.22% | 16.8 | 13.96-20.29 | 3,014.5 | 929-5,740 |
| Zoetermeer | 124.4 | 0.75% | 0.85% | 22.6 | 20.04-25.59 | 4,218.4 | 1,862-6,734 |
| Zoeterwoude | 8.1 | 0.05% | 0.05% | 21.9 | 15.91-29.18 | 3,994.4 | 1,727-6,887 |
| Zuidplas | 42.1 | 0.25% | 0.28% | 21.7 | 18.50-26.39 | 4,114.1 | 2,015-8,149 |
| Zundert | 21.4 | 0.13% | 0.13% | 19.3 | 15.47-27.22 | 3,749.7 | 1,774-6,493 |
| Zutphen | 47.0 | 0.28% | 0.24% | 17.0 | 13.44-21.95 | 2,891.6 | 954-5,497 |
| Zwartewaterland | 22.0 | 0.13% | 0.12% | 18.2 | 12.90-23.41 | 2,941.2 | 938-5,576 |
| Zwijndrecht | 44.3 | 0.27% | 0.27% | 20.4 | 16.95-24.71 | 3,663.0 | 1,708-6,487 |
| Zwolle | 126.6 | 0.76% | 0.70% | 18.2 | 15.20-22.12 | 2,761.9 | 889-5,058 |

Further, we also test the sensitivity of the transmission risk scores for the students demographic group to the time horizon by assigning all municipalities a rank based on their transmission risk scores and plotting these ranks at 17 and 21 days days as function of the ranks at 14 days in Fig. SI.7.

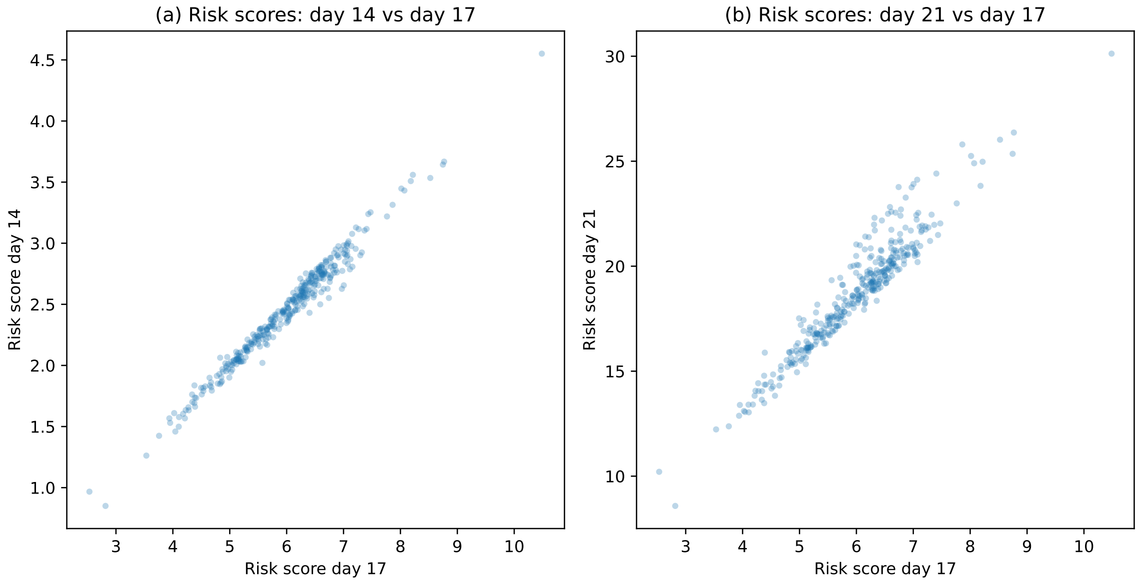


**Figure SI.7:** Scatter plots of transmission risk scores for the students demographic group between days 14, 17 and 21, for each municipality. Panels (a-b) show the risk scores are strongly correlated in their time horizons, indicated by the visual linearity of the scatter plots. Full risk score data can be found for each day in Tabs. SI.1-SI.3.

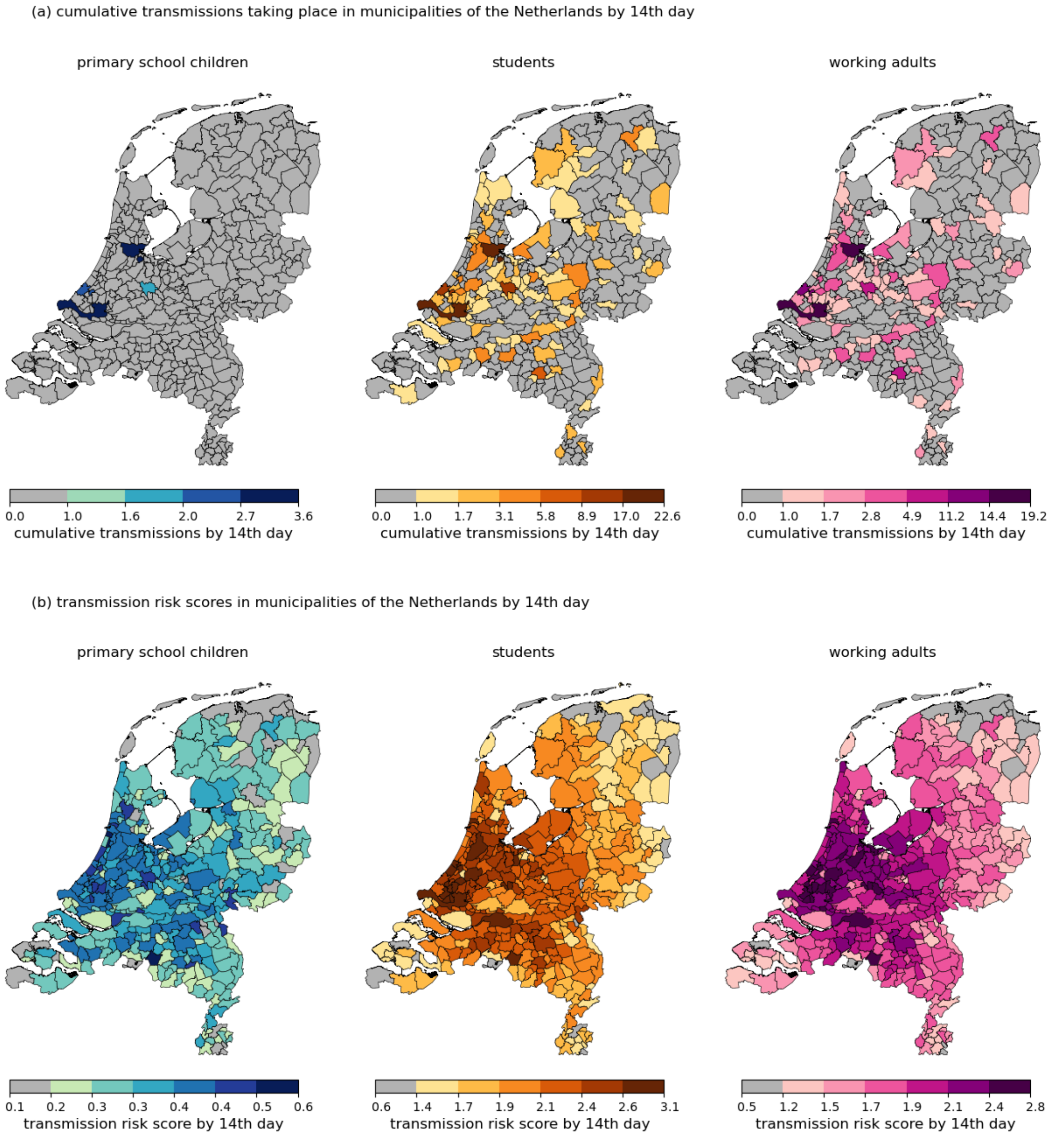


**Figure SI.8:** Municipality-resolved transmission maps, and transmission risk score maps using the 14th day, stratified by the demographic group into which the pathogen is introduced.

.

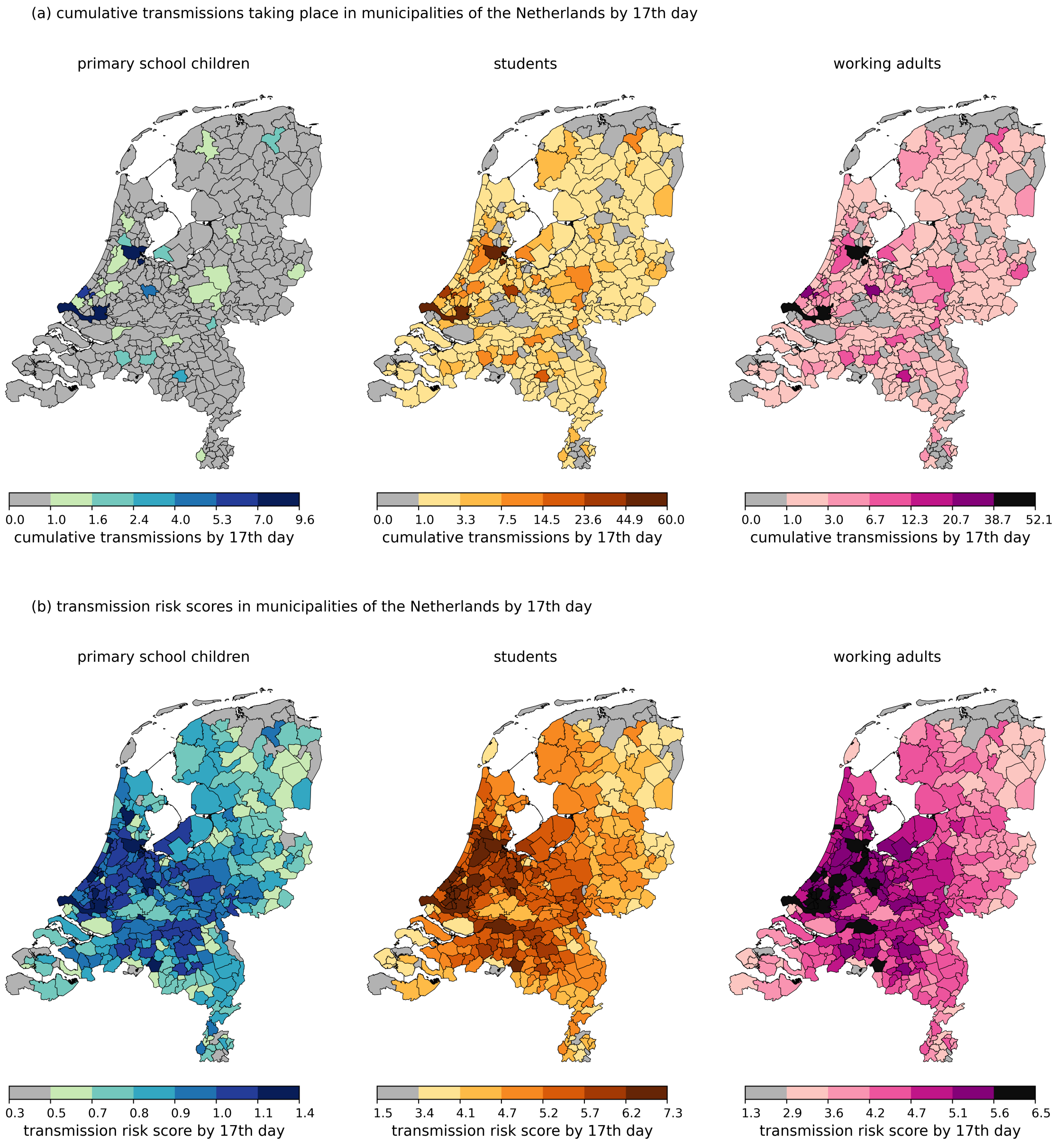


**Figure SI.9:** Municipality-resolved transmission maps, and transmission risk score maps using the 17th day, stratified by the demographic group into which the pathogen is introduced. This is the same Fig. 3 of the main text.

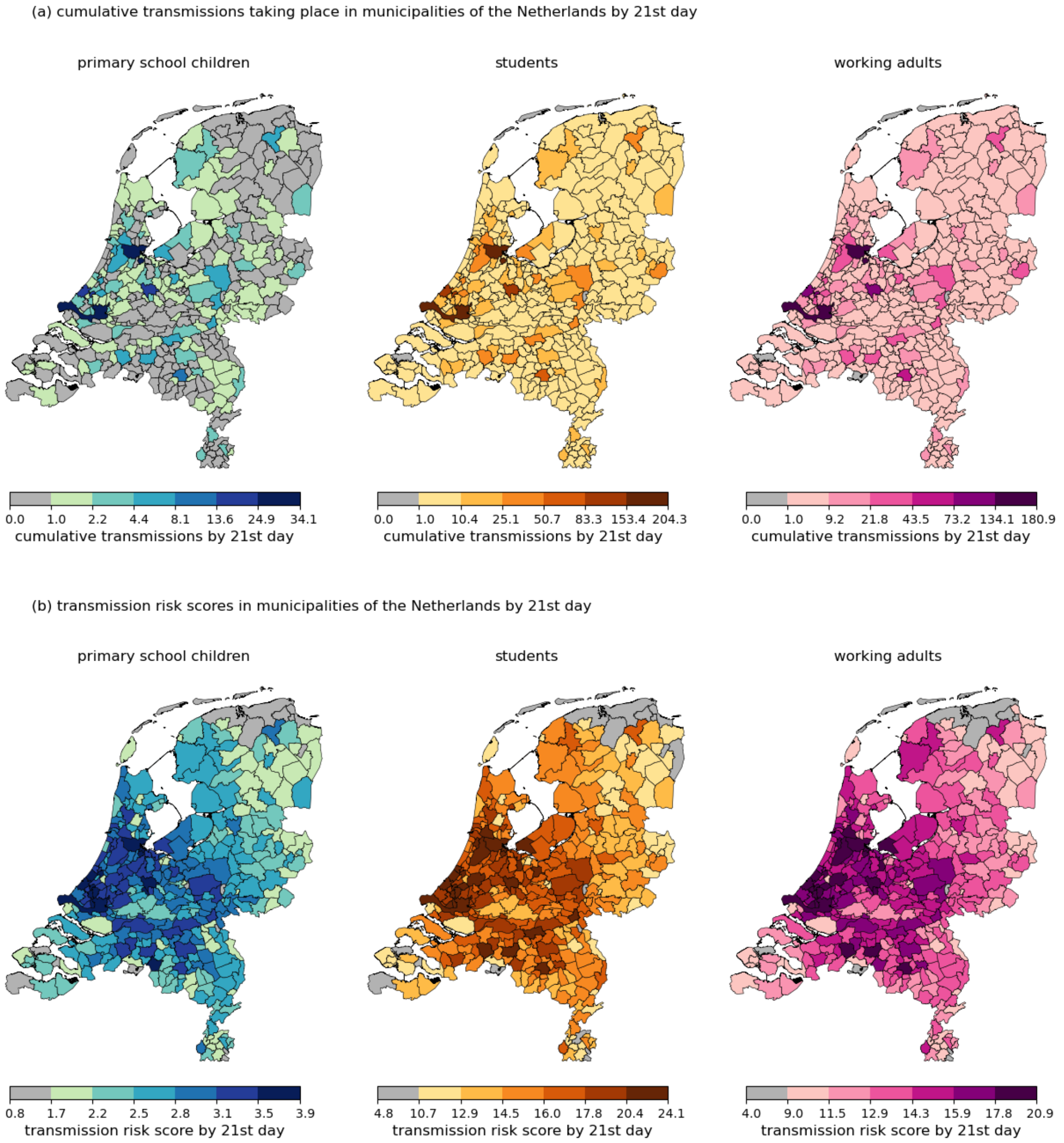


**Figure SI.10:** Municipality-resolved transmission maps, and transmission risk score maps using the 21st day, stratified by the demographic group into which the pathogen is introduced.

.

### SI D: Effect of stochasticity for the impact of behavioural changes and targeted interventions

Below in Fig. SI.11 we show the effect of stochasticity in terms of the 90% range for the simulation results in Fig. 4 of the main text.

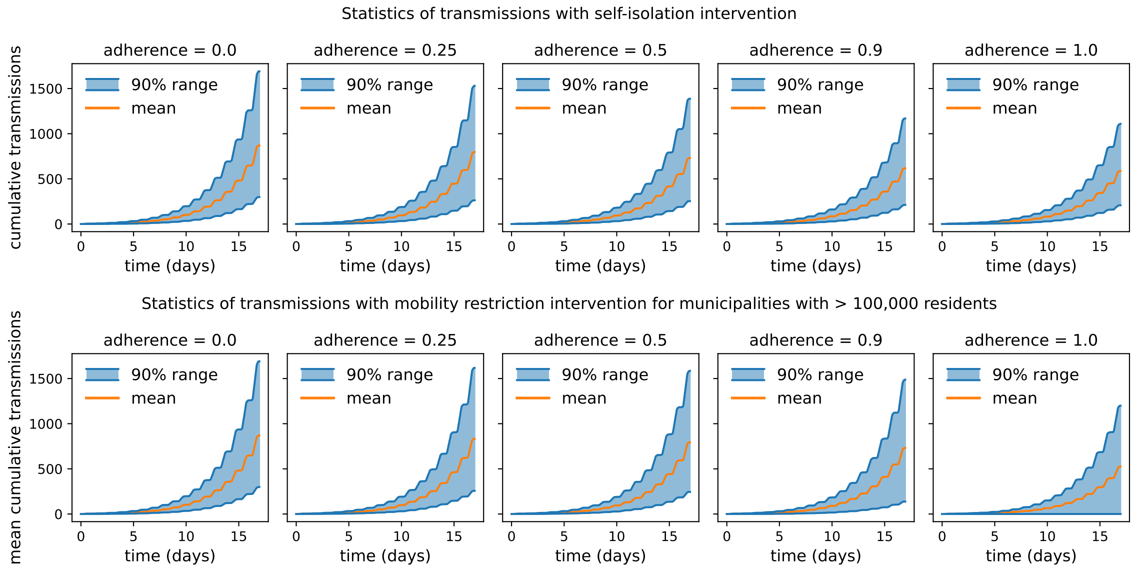


**Figure SI.11:** Effect of stochasticity — 90% ranges — for self-isolation (top row), and for targeted interventions (bottom row) for different adherence rates, adding error bars to the data in Fig. 4 of the main text. The data plotted are hourly, which explains the oscillating pattern due to the periodic day-night rhythm that impacts travel and mixing behaviour of actors.

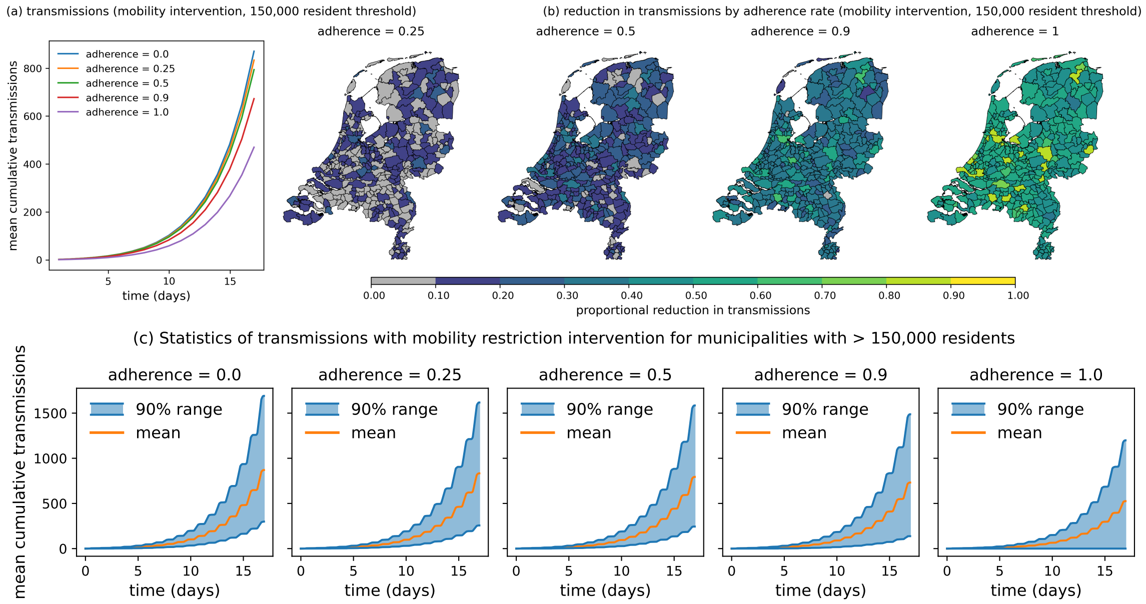


**Figure SI.12:** Reproduction of Fig. 4 of the main text for mobility restriction intervention scenario for a threshold of municipalities with 150,000 residents. Panel (a) shows transmissions over time for a period of 17 days. For all adherence rates we see similar to less reductions in Fig. 4 of the main text, which is to be expected. Reductions per municipality as shown in the maps of panel (b) show also show little effect of changing the threshold, other than the few cities that now no longer close their borders. Effect of stochasticity — 90% ranges — for the mobility restriction intervention with threshold at 150,000 residents (panel (c), for different adherence rates, adding error bars to the data in Fig. 4 of the main text. The data plotted are hourly, which explains the oscillating pattern due to the periodic day-night rhythm that impacts travel and mixing behaviour of actors.

### SI E: Sensitivity of mobility restriction intervention to municipality population size

In the mobility restriction intervention scenario in the main text, we chose a municipality population size of 100,000 as threshold to for municipalities to close down their borders. To test sensitivity of the measure to this threshold we reproduce panels (c) and (d) of Fig. 4 in the main text for a new threshold at a population size of 150,000 in Fig. SI.12. The effective change of this threshold is that now there are 17 cities with closed borders, where before there were 31. From the small difference between Fig. 4(c-d) of the main text and Fig. SI.12(a-b), we can conclude that most of the effect of the mobility restriction intervention scenario is caused by closing the borders of the 17 most populous municipalities.

### SI F: Calculation of $\mathcal{R}_0$

Below we outline how $\mathcal{R}_0$ has been calculated. Let $g(a)$ denote the generation-interval density, i.e. the probability density that a secondary infection occurs at infection age $a$ of the infector. Under early exponential growth $I(t) \propto e^{rt}$ with intrinsic growth rate $r$, the renewal equation implies the Euler–Lotka relation

$$1 \; = \; \mathcal{R}_0 \int_0^\infty e^{-ra}\, g(a)\, \mathrm{d}a,$$

so that

$$\mathcal{R}_0 \; = \; \frac{1}{M(-r)}, \qquad M(-r) \; = \; \int_0^\infty e^{-ra}\, g(a)\, \mathrm{d}a, \tag{SI.1}$$

where $M(-r)$ is the Laplace transform of the generation-interval distribution [1, 2].

In our model, the latent and infectious periods are stochastic variables represented by $T_E$ and $T_I$, respectively. Assuming independence and constant infectiousness during the infectious period, the infectiousness profile of an individual infected at time 0 is constant on the interval $[T_E,\, T_E + T_I)$ and zero elsewhere. Hence, the generation-interval density function is

$$g(a) \; = \; \frac{\mathbb{P}\big(T_E \le a < T_E + T_I\big)}{\mathbb{E}[T_I]}.$$

Substituting this into (SI.1) yields [1]

$$\mathcal{R}_0 \; = \; \frac{r\,\mathbb{E}[T_I]}{\mathbb{E}\big[e^{-rT_E}\big(1 - e^{-rT_I}\big)\big]} \; = \; \frac{r\,\mathbb{E}[T_I]}{\mathbb{E}[e^{-rT_E}]\,\Big(1 - \mathbb{E}[e^{-rT_I}]\Big)}, \tag{SI.2}$$

where the last identity follows from the independence assumption. Next, Eq (SI.2) is used to relate the epidemic growth rate $r$ to $\mathcal{R}_0$ for our choice of latent and infectious period distributions.

In the simulations, the latent and infectious periods are modelled as Weibull random variables, $T_E \sim \mathrm{Weibull}(k_E, \lambda_E)$ and $T_I \sim \mathrm{Weibull}(k_I, \lambda_I)$, with shape and scale parameters chosen to match a mean latent period of approximately 2 days and a mean infectious period of approximately 5 days: $k_E = 3.67$, $\lambda_E = 2.22$, $k_I = 2.39$, $\lambda_I = 5.64$. We estimated the intrinsic growth rate $r$ from early epidemic simulations by fitting an exponential function $I(t) \propto e^{rt}$ to the initial phase of the incidence curve. For the baseline parameters, this yielded $r \approx 0.295\,\mathrm{day}^{-1}$. Substituting these values into (SI.2) and solving numerically gives $\mathcal{R}_0 \; \approx \; 3.6$.

### References


[1] P. Yan, *Mathematical Epidemiology*, F. Brauer, P. van den Driessche, J. Wu, eds. (Springer, Berlin, 2008), vol. 1945 of *Lecture Notes in Mathematics*, pp. 229–293.

[2] O. Diekmann, H. Heesterbeek, T. Britton, *Mathematical Tools for Understanding Infectious Disease Dynamics* (Princeton University Press, Princeton, NJ, 2013).
